# Hemoglobin Interacts with Endothelial Nitric Oxide Synthase to Regulate Vasodilation in Human Resistance Arteries

**DOI:** 10.1101/2021.04.06.21255004

**Authors:** Steven D. Brooks, Olena Kamenyeva, Sundar Ganesan, Xianke Zeng, Rachel Smith, Dongying Ma, Juraj Kabat, Phillip Cruz, Brant Isakson, A. Parker Ruhl, Jeremy L. Davis, Hans C. Ackerman

## Abstract

**Background:** In small arteries, constriction of vascular smooth muscle triggers local release of nitric oxide from the adjacent endothelial cell. This feedback vasodilation is a homeostatic mechanism that opposes vasoconstriction. Here, we investigate the role of endothelial alpha globin as a regulator of directed nitric oxide signaling across the myoendothelial junction.

**Methods:** Human omental arteries 100-200µm in diameter were microdissected from omentum samples obtained during clinically indicated abdominal operations on NIH protocol 13-C-0176 (NCT01915225). Each artery was cannulated, perfused free of blood, and preserved for analysis or subjected to pressure myography. Preserved arteries underwent RNA extraction for gene expression; protein extraction for co-immunoprecipitation and Western blot; or immunostaining for multiphoton microscopy. Bio-layer interferometry quantified the binding of alpha globin to endothelial nitric oxide synthase (eNOS). *Ex vivo* pressure myography characterized arterial vasoreactivity before and after disruption of eNOS-Hb binding with an alpha globin mimetic peptide.

**Results:** *HBA1, HBA2, HBB*, and *NOS3* transcripts were abundant in RNA from the artery wall, and the blood cell gene *SLC4A1* was not. Beta globin and eNOS co-immunoprecipitated with alpha globin in protein extracted from human omental artery segments, suggesting an eNOS-hemoglobin complex. Biolayer interferometry studies estimated alpha globin to bind to the oxidase domain of eNOS with an equilibrium dissociation constant of 1.3 × 10^−6^ M.

Multiphoton microscopy of intact arteries revealed alpha globin, beta globin, and eNOS to co-localize within distinct punctates in a plane defined by the internal elastic lamina that separates endothelial cells from vascular smooth muscle. Förster resonance energy transfer confirmed close physical proximity of alpha globin to eNOS in situ.

Omental arteries constricted to 39.1 ± 3.2 % of baseline diameter in response to phenylephrine. After treatment with an alpha globin mimetic peptide, the same arteries constricted to 64.6 ± 1.6% of baseline (p < 0.01). Inhibition of NOS with L-NAME restored vasoconstriction in the mimetic peptide-treated arteries to 41.9 ± 2.0% (p < 0.0001).

**Conclusion:** Alpha globin and beta globin are expressed in the endothelium of human resistance arteries, form a complex with eNOS at the myoendothelial junction, and limit the release of nitric oxide triggered by alpha-1-adrenergic stimulation.

Graphical Abstract. Hemoglobin binds to endothelial NOS in the myoendothelial junction where it regulates the diffusion of nitric oxide that is produced in response to alpha-1-adrenergic signaling.
Phenylephrine (PE) engages alpha-1-adrenergic receptors (α_1_) on vascular smooth muscle cells triggering an influx of calcium (Ca^++^) and activating myosin light chain kinase (MLCK) to constrict muscle fibers (≈) and constrict the artery. Calcium enters the endothelial cell via putative gap junctions where it activates endothelial nitric oxide synthase (NOS) via calmodulin (CM). Nitric oxide (NO) produced by NOS can diffuse into the smooth muscle cell where it activates myosin light chain phosphatase (MLCP) via soluble guanylate cyclase and cGMP (both not shown) to relax smooth muscle fibers and dilate the artery. Hemoglobin (αβαβ) bound to NOS prevents the diffusion of NO into the smooth muscle cell likely by catalyzing the reaction with oxygen (O_2_) to produce nitrate (NO_3-_), an anion that cannot diffuse across the cell membrane. When hemoglobin is displaced from NOS with a mimetic peptide (not shown), NO diffusion increases and counteracts vasoconstriction; when NOS is inhibited by L-NAME, the mimetic peptide has no effect. Thus hemoglobin limits the diffusion of NO across the myoendothelial junction in the setting of alpha-1-adrenergic stimulation.

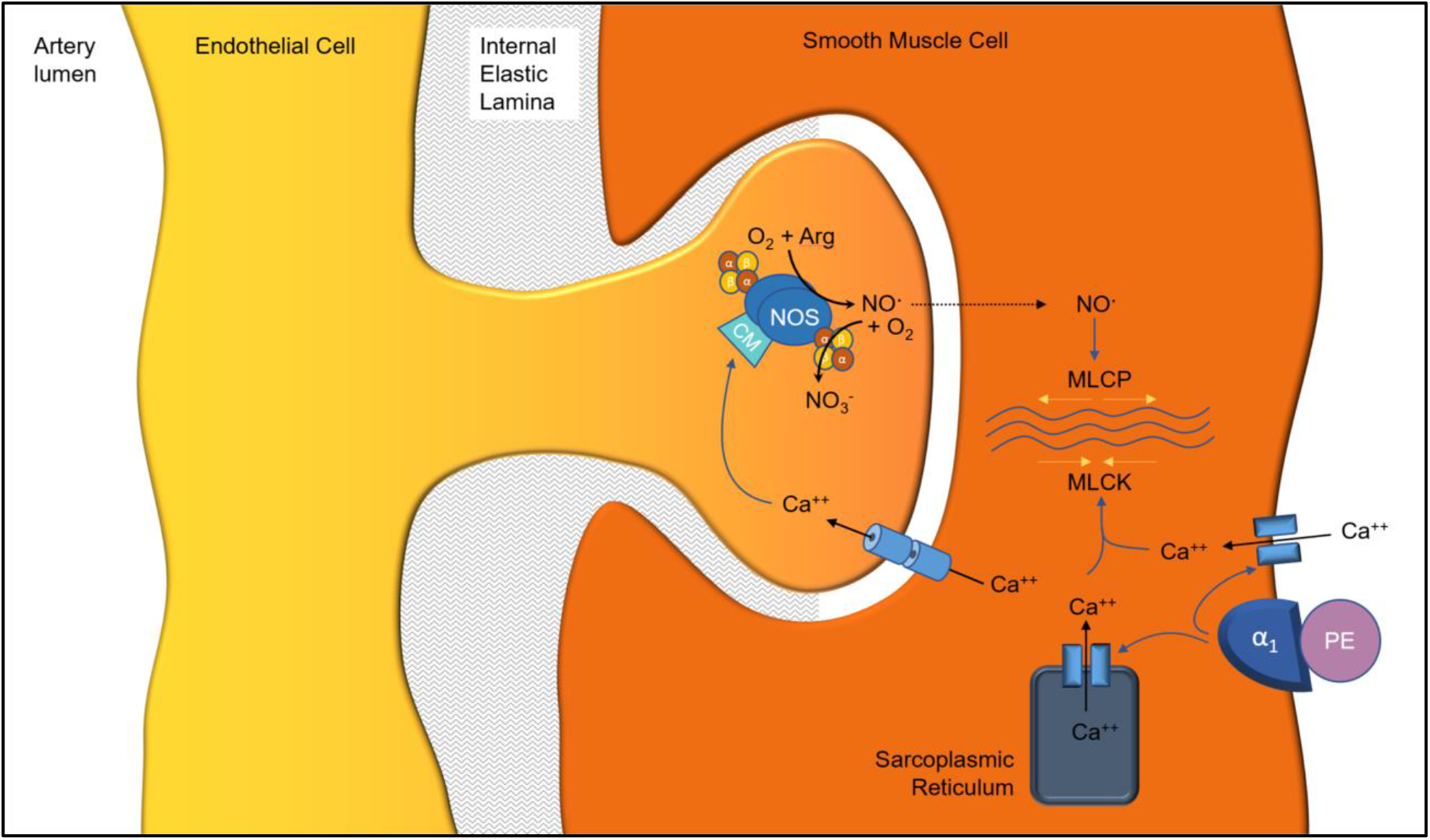

## INTRODUCTION

Peripheral vascular resistance is primarily provided by small arteries of the vascular system that link conduit arteries to arterioles feeding capillary beds.^1^ Peripheral small arteries can constrict to increase blood pressure or redistribute blood flow to meet regional demands.^2–4^ Third and fourth order arteries that perfuse the visceral organs are especially dynamic in their vasoreactivity.^5–7^ These arteries can dilate to provide increased blood flow during digestion^2^ and constrict to divert blood flow away from visceral organs to maintain cerebral pressure or to support skeletal muscle during physical activity.^8,9^ The signals that mediate vasoreactivity in small mesenteric arteries can act through vascular endothelial cells or directly on vascular smooth muscle cells through nervous innervation.^10–12^

Alpha-adrenergic signaling plays a major role in the regulation of systemic blood pressure by directly stimulating vascular smooth muscle cells to constrict.^13^ Activation of alpha-1-adrenergic receptors triggers vascular smooth muscle cells (vSMCs) to constrict by activating PL-C and increasing IP3, leading to increased intracellular calcium release and subsequent activation of myosin light chain kinse.^14–17^ This vasoconstrictive stimulus is counterbalanced by a vasodilatory signal emanating from the adjacent endothelial cell, which produces both nitric oxide (NO)^18^ and endothelial-derived hyperpolarizing factor (EDHF) in response to adrenergic constriction of vSMCs.^19^ The depolarization of vSMCs during adrenergic stimulation opens L-type voltage gated calcium channels on their membrane,^20^ near the myoendothelial junctions (MEJs) that connect vSMCs with endothelial cells.^21^ The local influx of calcium near the MEJ within the vSMC is then communicated to the endothelial cell via both direct and indirect calcium and IP_3_ signaling.^21–24^ This signaling from the vSMC to the endothelium induces intracellular release of calcium within the endothelium to produce vasodilatory stimuli to relax the vascular smooth muscle back to its resting state.^18,21,25,26^ These vasodilatory stimuli include the efflux of K^+^ ions from the endothelial cell that hyperpolarize the vSMC^19,21,26^ and calcium-dependent activation of eNOS via calmodulin to produce NO, a potent vasodilatory molecule.^18,19,27,28^ Thus, alpha-1-adrenergic vasoconstriction induces a counterbalancing vasodilation signal from endothelium, ^18,21,28^ a mechanism referred to as feedback vasodilation.^21^

The release of NO from eNOS across the myoendothelial junction is under a final level of regulation through protein-protein interactions of eNOS with alpha globin. Important studies of skeletal muscle resistance arteries from mice demonstrated that alpha globin is found in vascular endothelial cells where it binds to eNOS and regulates the diffusion of nitric oxide across the MEJ during endothelium-dependent vasodilation and in response to alpha-1-adrenergic vasoconstriction.^29,30^ In mice, formation of the alpha globin-eNOS complex requires the presence of alpha hemoglobin stabilizing protein (AHSP).^31^ Molecular modeling studies identified a 10 amino acid sequence, conserved across species, that was predicted to interact with eNOS.^30,32^ This sequence was used to encode an alpha globin mimetic peptide, named HbaX, which was shown to disrupt the association of alpha globin with eNOS *in vivo*, resulting in increased NO bioavailability, reduced alpha-1-adrenergic vasoconstriction, lower mean arterial blood pressure, and protection against angiotensin-2 induced hypertension in mice.^30,32^

Given the potential importance of this novel vasoregulatory pathway in the human circulatory system, we sought to determine the expression, sub-cellular localization, and binding partners of alpha globin in human omental resistance arteries. Then, we used the HbaX alpha globin mimetic peptide to disrupt the association of alpha globin with eNOS in human omental arteries to characterize the functional role of alpha globin in regulating vasoreactivity. Together, these studies establish a role for endothelial alpha globin plus beta globin (i.e., hemoglobin) as a restrictor of nitric oxide diffusion in human omental arteries that directly modulates feedback vasodilation to an alpha-1-adrenergic agonist.

## METHODS

### Collection of omental arteries

Human omental tissue was collected from patients during clinically indicated abdominal operations at the NIH Clinical Center. All patients provided informed consent for tissue procurement on IRB-approved protocol 13-C-0176 (NCT01915225). All tissue removed by the surgeon was immediately submerged in cold 4 °C Krebs-HEPES (KH) buffer (pH 7.4) and kept on ice during dissection and artery isolation. Small arteries were dissected away from the tissue, cannulated on one end with a glass micropipette, and gently perfused with cold KH buffer to remove RBCs from the vessel lumen. Arteries were then prepared for downstream application.

### Gene expression studies

Gene expression was measured by reverse-transcriptase droplet digital PCR (ddPCR) using primer/probe assays for *HBA1, HBA2, HBB, NOS3*, and *SLC4A1*(Bio-Rad) in arteries from human omental tissue and human subcutaneous adipose tissue in RNAlater, and human whole blood in PaxGene RNA tubes. Gene expression was quantified as total transcripts of target gene per 1 ng of cDNA.

### Co-Immunoprecipitation and Western blot

Protein was extracted (Minute™ Total Protein Extraction Kit for Blood Vessels, InventBiotech # SA-03-BV) from perfused small omental arteries from nine individual donors (30-50 arteries each). For Western blot, 20 µg of total blood vessels proteins were separated by SDS-PAGE and immunoblotted with anti-Band 3 polyclonal antibody (1:1000, Fisher Scientific #PA5-80030). HRP signal was detected by enhanced chemiluminescence (Pierce).

Co-immunoprecipitation of alpha globin, beta globin, and eNOS was performed on 600 µg of blood vessels lysates pooled from two individual donors. Immunoprecipitation (IP) was performed with alpha globin polyclonal antibody (HBA, 1:1000, Proteintech, #14537-1-AP) or normal rabbit IgG antibody (Cell signaling #2729). Proteins were bound to magnetic beads and pelleted by a magnet stand. Samples were then analyzed by SDS-PAGE and Western blot with the IP antibody, beta globin polyclonal antibody (1:1000, Proteintech # 16216-1-AP), eNOS polyclonal antibody (1:1000, Cell Signaling #32027), and cytochrome B5 reductase 3 (Cyb5R3) monoclonal antibody (Abcam ab133247).

### Protein-protein binding affinity assessed by biolayer interferometry

Biolayer interferometry experiments were performed on the automated eight-channel Octet RED96 instrument (ForteBio). Alpha globin was biotinylated using EZ-Link™ NHS-PEG4 Biotinylation Kit (Thermofisher Scientific) and immobilized onto Streptavidin (SA) Biosensors. Binding kinetics and data traces were obtained using Data Analysis software v8.2 (ForteBio).

### Molecular modeling of alpha globin, hemoglobin, eNOS, and cytochrome B5 reductase

Molecular modeling, graphics, and analysis were produced using UCSF Chimera package^33^ and the virtual reality mode of UCSF ChimeraX.^34^ Protein-protein docking was performed using the HADDOCK 2.4 online server.^35^ Computational docking of eNOS to alpha globin performed using HADDOCK was guided by specifying the residues from the HbaX peptide on alpha globin and the general area of eNOS that contacted alpha globin on the original model.^30^

### Antibody labeling of intact omental arteries

Isolated and perfused small omental arteries were fixed in 4% formaldehyde in PBS solution (ImageIT, Invitrogen). Vessels undergoing immunofluorescent labeling were permeabilized with 0.1% Triton-X 100 and incubated overnight at 4°C with primary conjugated antibodies for alpha globin (Abcam ab215919), beta globin (SantaCruz, sc-21757-AF546), or eNOS (SantaCruz, sc-376751-AF594). DAPI (Invitrogen) labeling was performed to demark cell nuclei. Vessels undergoing immunofluorescent labeling with multiple antibodies were permeabilized, blocked in goat serum, and incubated overnight at 4°C with primary antibodies for alpha globin (Abcam ab92492), beta globin (Santa Cruz sc-21757), and/or eNOS (Abcam ab76198). Arteries were then washed and stained with secondary fluorescent antibodies.

### Multiphoton imaging of intact omental arteries

Images were acquired using a Leica SP8 Inverted DIVE (Deep In Vivo Explorer) multiphoton (MP) microscope (Leica Microsystems, Buffalo Grove, IL) with 25.0X and 40.0X Water Immersion Objective, as previously described.^36^ Multiphoton excitation was performed at 880nm (MaiTai DeepSee, Spectra Physics) and 1150nm (InSight DeepSee, Spectra Physics) and emitted fluorescence was measured using a 4Tune 4 HyD (4 tunable non-descanned hybrid detectors (HyDs)) reflected light detector. Multiphoton images were collected as a Z-stack in 1-3µm steps for up to 50 single phases. The collected image files were exported into Huygens Pro SVI and Imaris Bitplane for image deconvolution, processing, and analysis.

### Fluorescence lifetime imaging microscopy (FLIM)

Images were acquired using the Leica SP8 Inverted DIVE multiphoton microscope. FLIM images were collected using multiphoton excitation with Mai Tai-MP laser (Spectra Physics) tuned at 880nm at a 80Mhz frequency and images were simultaneously acquired for MP imaging and FLIM using 4-Tune External Hybrid detectors. Images were acquired at 512-512-pixel format, collecting in excess of 2,000 photons per pixel. Fluorescent Lifetime Decays and Förster resonance energy transfer (FRET/FLIM) efficiency transients and FRET-FLIM images were collected, analyzed, and processed using LASX Single Molecule detection analysis software.

### Multiphoton image processing and analysis of fluorescence signal intensity

The Leica Image File (.lif) for each artery was deconvolved to increase resolution and decrease noise and background (Huygens Pro, SVI). Region of interests (ROI) were created and analyzed for signal intensity, density, and volume in Imaris (Bitplane). Mean fluorescence intensity in each detector channel was plotted for each surface object, as well object size in voxels. The density of globin complexes was defined as number of surface objects within the ROI.

### Alpha globin mimetic peptide

A previously published molecular modeling study identified a conserved 10 amino acid sequence LSFPTTKTYF that was predicted to facilitate binding of alpha globin to eNOS.^30^ This sequence was combined with an N-terminal HIV-*tat* tag sequence (YGRKKRRQRRR) to provide membrane permeability (YGRKKRRQRRRLSFPTTKTYF, Anaspec). This mimetic peptide, called HbaX, has been previously patented for its therapeutic potential. A scrambled version of the peptide with an N-terminal HIV-*tat* tag sequence (YGRKKRRQRRRFPYFSTKLTT, Anaspec) was used as a peptide treatment control.

### Isolated artery pressure myography

The pressure myography methodology utilized here for omental arteries is consistent with published protocols for assessing reactivity of mesenteric articles using a DMT pressure myograph.^37^ Arteries 100-200 µm in diameter were dissected from omental tissue on ice and transferred to the culture myograph wells (DMT-USA CM204); arterial inner diameter was measured by video microscope (DMT-USA) with digital calipers (MyoVIEW, DMT-USA).

All arteries used developed at least 20% myogenic tone after pressurization, consistent with standards for pressure myography.^37–39^ Arteries were assessed for vasoconstriction response to the alpha-1-adrenoreceptor agonist phenylephrine (PE) [10^−9^ - 10^−3^] M (Sigma-Aldrich). Dose response to PE was then assessed after incubation with either the alpha globin mimetic peptide HbaX or a control peptide (5 µmol/L in KH buffer), and again following incubation with the NOS inhibitor Nω-Nitro-L-arginine methyl ester HCl (L-NAME, 10^−4^ M)(Sigma-Aldrich).

Full descriptions of methodological details are provided in Appendix A (Supplemental Materials).

## RESULTS

### *HBA1, HBA2, HBB*, and *eNOS* are expressed in blood-free human resistance artery tissue

We used reverse-transcriptase droplet digital PCR (RT-ddPCR) to quantify transcripts from *HBA1, HBA2, HBB*, and *NOS3* in human omental arteries. All four genes were highly expressed in omental arteries (Fig. 1A) and subcutaneous arteries (Supplemental Fig. 1A). To assess for potential contamination of arterial tissue with residual blood, we checked for transcripts from an erythroid-specific gene, *SLC4A1. SLC4A1* transcripts were abundant in whole blood (geometric mean 4306 transcripts per ng RNA; 95% CI 899, 20638) but very low or absent in omental arteries (2.5; 95% CI 0, 5.2) and subcutaneous arteries (1.8; 95% CI 0, 5.3). Further evidence for artery-specific expression of the globin genes was provided by the unique expression ratios of *SLC4A1*/*HBA1* in arterial tissue versus blood (0.0018 ± 0.0011 vs 0.0224 ± 0.0087; p = 0.016; Fig. 1B). Notably, the ratio of *HBA1*/*HBA2* was distinct in arterial tissue compared to whole blood (0.60 ± 0.14 vs 0.12 ± 0.05; p = 0.01)(Fig. 1C), raising the possibility that *HBA1* and *HBA2* are under differential tissue-specific transcriptional control in the artery compared to in the red blood cell progenitor.

**Figure 1:**
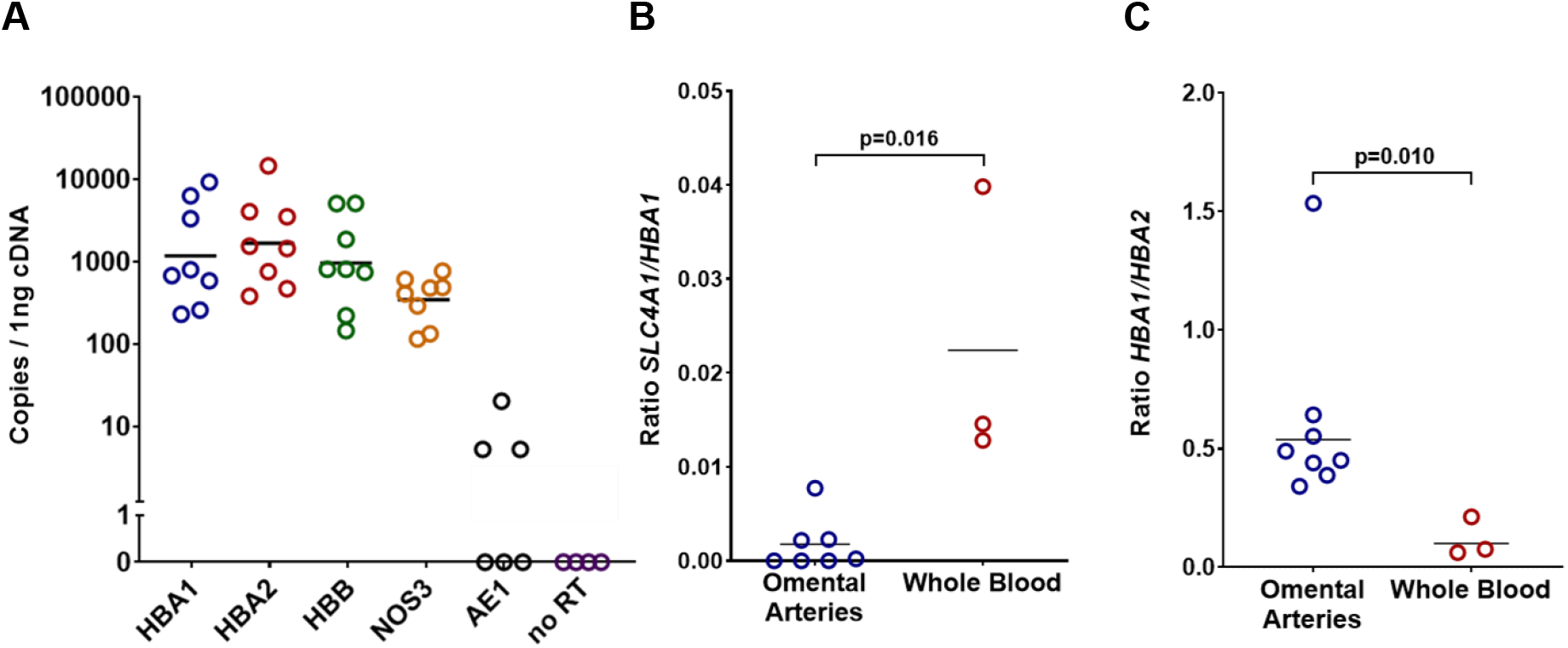
Gene expression in perfused human small arteries dissected from omental tissue (n=8); data is plotted with geometric mean. **A)** Total copies of *HBA1, HBA2, HBB, NOS3, SLC4A1* and a no-reverse transcriptase reaction control, per 1 ng of cDNA. **B)** The expression ratio of the erythrocyte specific marker *SLC4A1* to *HBA1* is lower in omental arteries (n=8) compared to human whole blood (n=3)(p=0.016), and **C)** the expression ratio *HBA1* to *HBA2* is lower in omental arteries (n=8) compared to human whole blood (n=3)(p=0.010).

### Hemoglobin forms a complex with eNOS in human resistance arteries

Based on the observation of FRET between antibodies labeling alpha globin and eNOS, we were interested in whether these proteins form a stable complex *in vivo*. Approximately 30-50 perfused, intact omentum arteries from each of six more donors were homogenized and protein was extracted. Immunoprecipitation was performed with an antibody against alpha globin. Western blot detected alpha globin, beta globin, and eNOS in the anti-alpha globin immunoprecipitate but not in a control immunoprecipitate (Fig. 2A). These results indicate that these three proteins not only co-localize within omental arteries, but also form a stable complex *in vivo*. To confirm there was no contamination of arterial lysate from residual erythrocytes, we performed Western blot on the lysate with an antibody for SLC4A1, which is the most abundant non-globin protein in the erythrocyte proteome.^40^ Even at maximal exposure, no SLC4A1 was detected in pooled arterial lysates (Fig. 2B). Based on previously published work that cytochrome B5 reductase 3 (Cyb5R3) functions as a hemoglobin reductase in rodent models,^29,41^ we probed for Cyb5R3 in the alpha globin immunopreciptate, and found that Cyb5R3 is present in perfused human artery lysates and bound to alpha globin.

**Figure 2:**
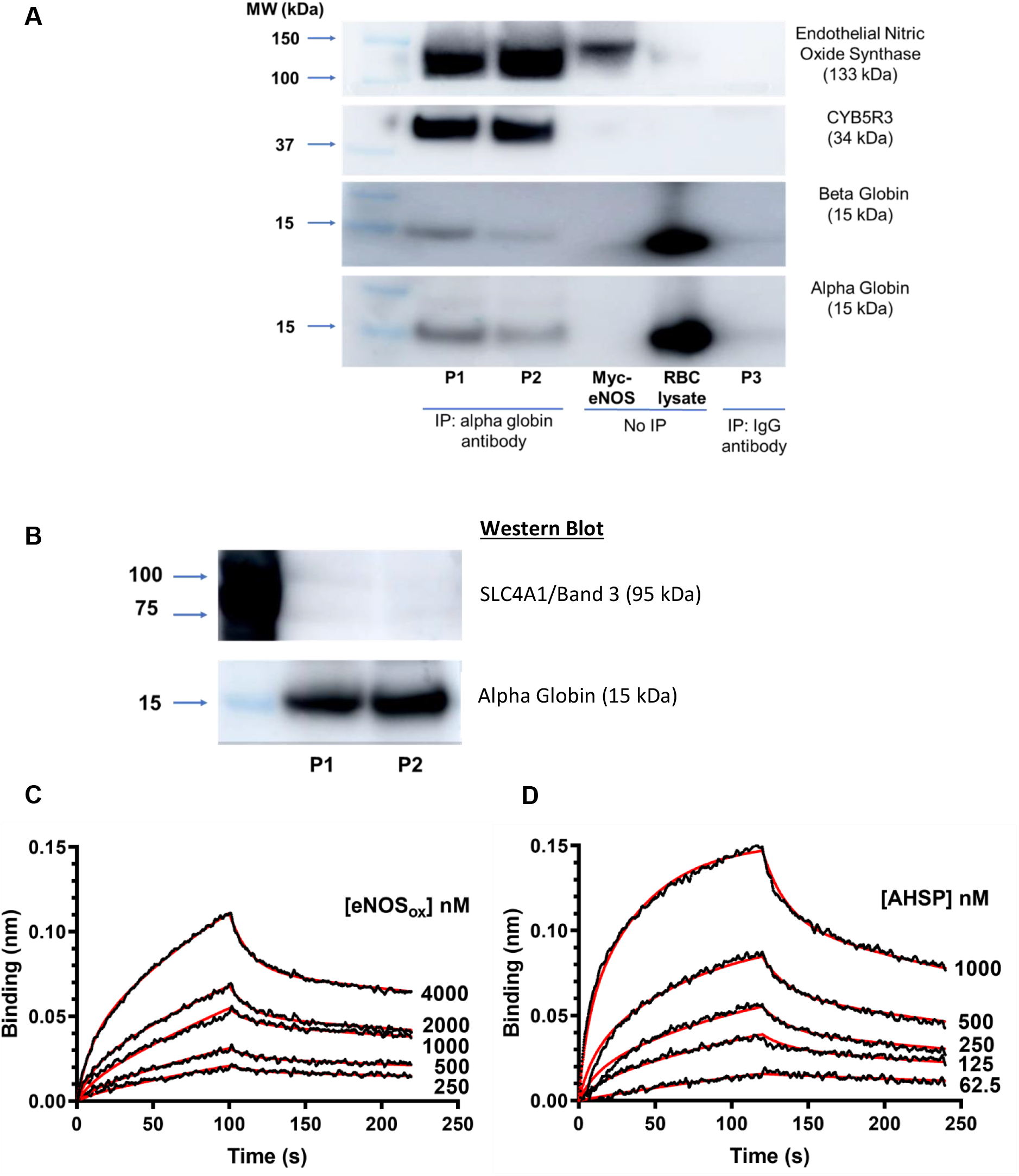
**A)** Beta globin, eNOS, and cytochrome B5 reductase 3 (Cyb5R3) co-immunoprecipitate with alpha globin in lysates of perfused human omental arteries. P1, P2, and P3 each represent an independent replicate consisting of pooled flash-frozen arteries from two donors (n=30 arteries from each donor). P1 and P2 were incubated with an alpha globin antibody to immunopreciptate alpha globin and bound protein partners. P3 was incubated instead with an IgG antibody as an negative control. Western blot was performed on the immunoprecipitated with antibodies for alpha globin, beta globin, Cyb5R3, or eNOS. Myc-eNOS purified from a cell overexpression lysate served as a positive control. RBC lysate served as a positive control for alpha and beta globin. **B)** Band 3/SLC4A1 is not detected in protein lysates from pooled human omental arteries. Prior to immunoprecipitation, whole protein lysate was loaded into a gel and probed by Western Blot for SLC4A1 and alpha globin. Gel exposure was maximized to probe for any SLC4A1 signal in pooled samples P1 or P2; while alpha globin was abundant in both lysates, no SLC4A1 was detected, indicating that arteries were fully perfused of RBCs. **C-D)** Sensorgram Traces for eNOS and AHSP with alpha-globin showing association and dissociation; raw datasets shown in black, fitting curves shown in red. **C)** Binding between Alpha-globin and eNOS_ox_. The sensors were dipped in 200 mL of eNOS_ox_ solution at 4000 nM, 2000 nM, 1000 nM, 500 nM, 250 nM for 120 seconds for association, and then moved to assay buffer for another 120 seconds for dissociation. **D)** Binding between Alpha-globin and AHSP: The sensors were dipped in 200 mL of AHSP solution at 1000 nM, 500 nM, 250 nM, 125 nM, 62.5 nM for 120 seconds for association, and then moved to assay buffer for another 120 seconds for dissociation.

Modeling studies had previously predicted alpha globin to interact with the oxidase domain of eNOS.^30^ To test this prediction experimentally, we measured the binding of recombinant human eNOS oxidase domain (eNOS_ox_) to purified human alpha globin using bio-layer interferometry (Fig. 2C). eNOS_ox_ bound to immobilized alpha globin with moderate affinity (K_D_ = 1.31 × 10^−6^ ± 1.35 × 10^−7^ M; Fig. 2C). For comparison, alpha hemoglobin stabilizing protein bound to alpha globin with stronger affinity (K_D_ = 1.09 × 10^−7^ ± 6.50 × 10^−9^ M; Fig. 2D). This biophysical analysis of the binding of alpha globin to eNOS corroborates the FRET and co-immunoprecipitation data supporting the observation of an eNOS-hemoglobin complex.

### Alpha globin and beta globin co-localize with eNOS at the myoendothelial junctions of human omental arteries

To examine the sub-cellular localization of alpha and beta globin proteins with eNOS in resistance arteries, we performed immunofluorescent multiphoton microscopy on intact human omental artery segments. Multiphoton imaging revealed distinct punctates distributed in the wall of the artery (Fig. 3A) that exhibited broad autofluorescent properties (Supplemental Figure 2 A-F). The broad autofluorescence of these punctates is similar to the those of hemoglobin under MP excitation^42^ (Supplemental Fig. 3, A-D). We compared the autofluorescent properties of the arterial wall punctates against the autofluorescence of hemoglobin within intact red blood cells (RBCs) in the same field of view (Fig. 3B), and discovered the arterial wall punctates to be distinct from the RBCs; they were smaller in volume and had greater signal intensities than the RBCs across all channels (Supplemental Figure 3, E-H).

**Figure 3:**
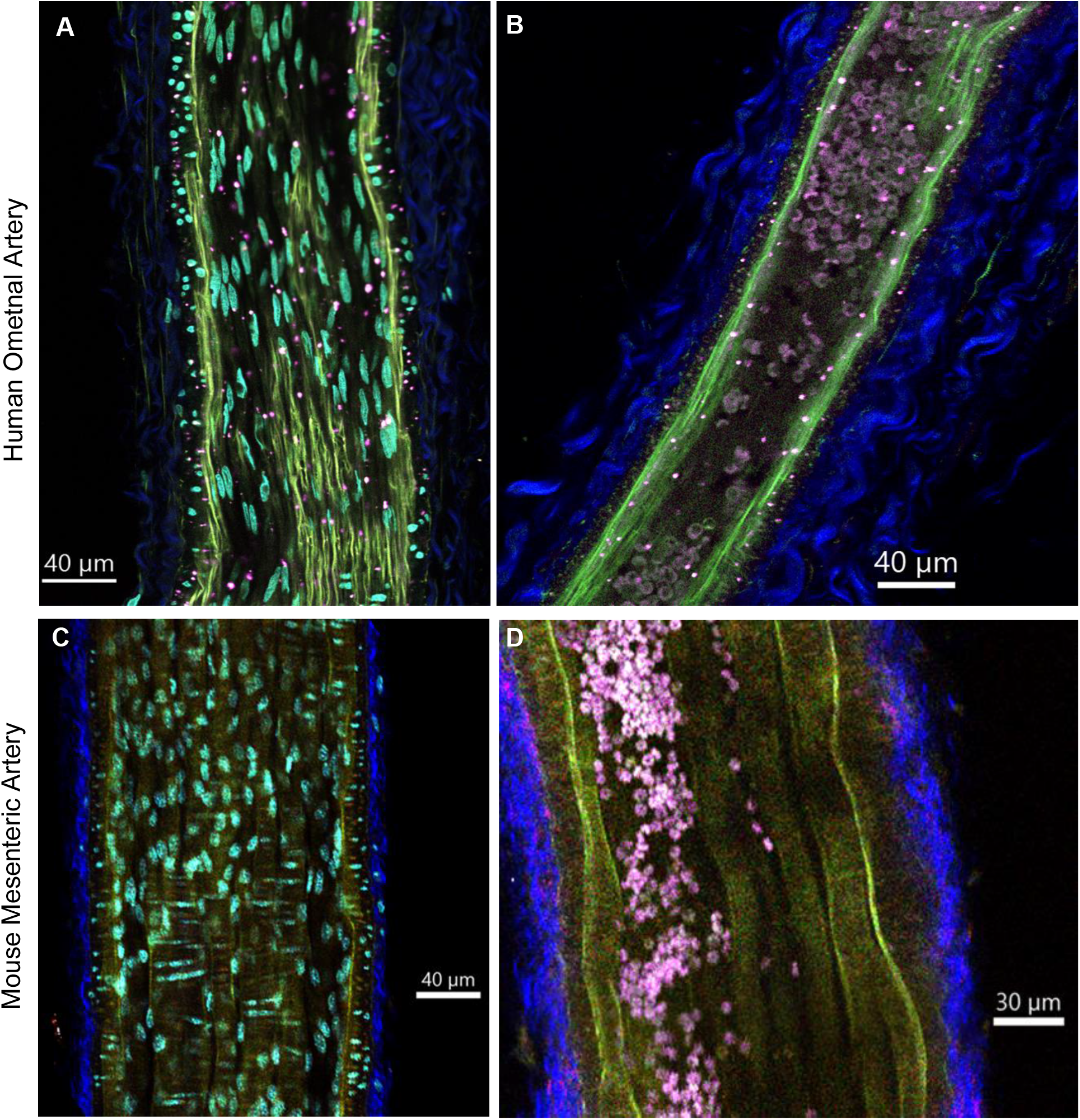
Hemoglobin presents as autofluorescent punctates in the walls of omental arteries. **A)** Longitudinal view of intact omental artery viewed from a 1 µm Z-step in plane with the arterial endothelium and internal elastic lamina (IEL, green). The hemoglobin-eNOS complexes (bright pink punctates) are broadly autofluorescent. Endothelial cell nuclei (DAPI, light blue) are aligned in the direction of flow within the vessel. The outer collagen layer demonstrates second harmonic generation (SHG, blue). **B)** Longitudinal view of intact omental artery viewed from a 1 µm Z-step in plane with the arterial endothelium and internal elastic lamina (IEL, green) in a non-perfused artery; residual RBCs (bright pink) demonstrate broad autofluorescence. **C)** Longitudinal view of intact perfused murine mesenteric artery viewed from a 1 µm Z-step in plane with the arterial endothelium and the IEL (green); endothelial nuclei (light blue) are visible. **D)** View of non-perfused intact murine mesenteric artery; residual RBCs (bright pink) demonstrate broad autofluorescence.

We then imaged a mesenteric artery from a mouse, which had been perfused, fixed, and imaged using the same methodology, to determine whether eNOS-alpha globin complexes in murine arteries would exhibit similar autofluorescence despite only expressing alpha globin and not beta globin in the arterial wall. Within the murine mesenteric artery, we did not observe any autofluorescent punctates in the arterial wall (Fig. 3C); however, the internal elastic lamina (IEL) was autofluorescent in a similar pattern to human omental arteries, and the outer collagen layer had a similar second harmonic generation (SHG). We then imaged a murine mesenteric artery that had not been fully perfused of RBCs to see if murine hemoglobin within red cells was autofluorescent (Fig. 3D). The murine RBCs within the mesenteric artery demonstrated similar autofluorescence to human RBCs within an omental artery, while no autofluorescent punctates were visible within the arterial wall.

To investigate the protein constituents of this autofluorescent complex, we used the unique fluorescence lifetimes, rather than the wavelengths, of the antibody-conjugated fluorophores to distinguish the autofluorescent punctate from antibodies binding individually to each of alpha globin (Fig. 4 A-C), beta globin (Fig. 4 D-F), and eNOS (Fig. 4, G-I). The close physical proximity of alpha globin and eNOS was further evaluated by co-immunostaining alpha globin and eNOS with fluorophore conjugated antibodies and assessing for FRET (Fig. 4 J, K). The fluorescence lifetime of singly labeled alpha globin (3.95 ± 0.04 ns) and eNOS (3.96 ± 0.02 ns) shifted to a shorter lifetime when co-immunostained (3.29 ± 0.05 ns; p < 0.0001), indicating a FRET effect that occurs when two fluorophores are within 10 Angstroms of each other. Together, these observations were consistent with a multi-protein complex that contains alpha globin, beta globin, and eNOS, which comprises the broadly autofluorescent punctate visible in the arterial wall.

**Figure 4:**
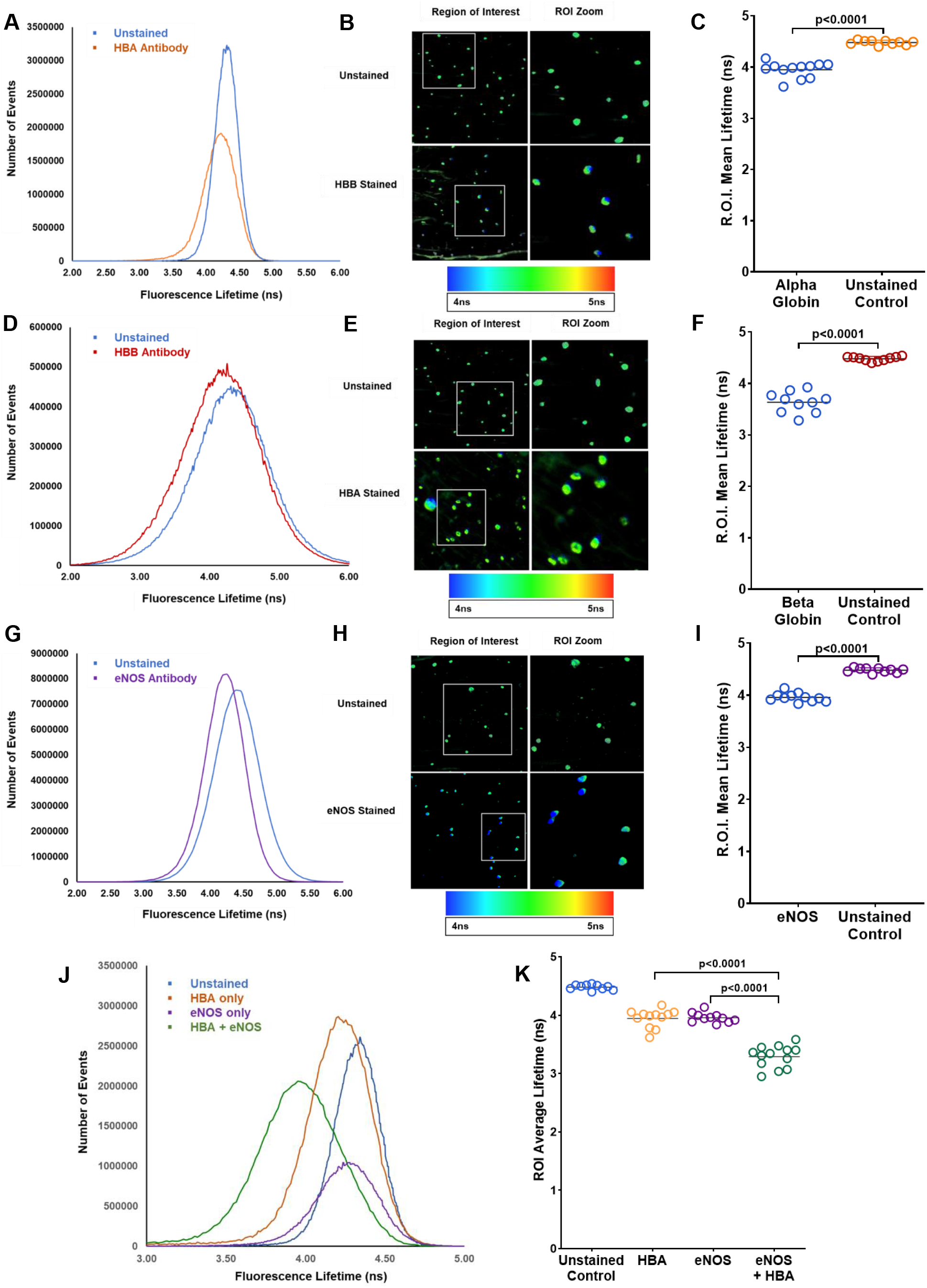
Fluorescence Lifetime Imaging Microscopy (FLIM) of intact omental arteries shows specific binding of antibodies at the autofluorescent punctate. Arteries were split, with one half imaged unstained and the other half labeled with a specific antibody. Total number of fluorescent lifetime events were plotted for the unstained and antibody labeled sections for **A)** alpha globin, **D)** beta globin, and **G)** eNOS. To focus on fluorescent lifetime differences within the distinct autofluorescent punctates with and without antibody labeling, Regions of Interest (ROIs) were selected for 10 of the autofluorescent antibody complexes from each labeling condition **(B, E, I)**; the lifetime pseudocolor lookup table encodes lower lifetimes as blue and longer lifetimes as green or red. The mean fluorescent lifetime within each ROI was then calculated and compared to unstained control **(C, F, I)**. Mean fluorescence was significantly decreased following labeling with alpha globin **(C)**, beta globin **(F)**, or eNOS **(I)** (line shown at geometric mean; p<0.0001 for all comparisons). **J)** FLIM was then performed on an intact omental artery as unstained, labeled with alpha globin antibody only, labeled with eNOS antibody only, or co-labeled with antibodies for alpha globin and eNOS. Co-labeling with eNOS and alpha globin antibodies shifted the peak lifetime further left of alpha globin or eNOS alone, indicating that the two antibodies were generating a possible FRET-like effect due to spatial proximity. **K)** ROIs were selected for each labeling condition as mean fluorescent lifetimes calculated for each ROI. Alpha globin antibody and eNOS antibody each decreased mean lifetime compared to unstained control (p<0.0001); co-labeling with both eNOS and alpha globin antibodies further decreased the mean fluorescence lifetime compared to single-stained arteries (p<0.0001).

To identify the localization of the hemoglobin-eNOS complexes, whole intact arteries were imaged by multiphoton microscopy (Fig. 5A, Supplemental Video 1). The eNOS-hemoglobin complexes localized to the plane of the internal elastic lamina (IEL) as seen in a transverse view (Fig. 5B). They are in closer proximity to the DAPI-stained endothelial cell nuclei (oriented in the direction of flow) than to the vascular smooth muscle nuclei (oriented perpendicular to flow). A longitudinal view reveals the eNOS-hemoglobin complexes to be in the same 1µm plane as the IEL (Fig. 5C); viewing the artery in three dimensions, the pink punctates are visible protruding through and on the endothelial side of the IEL (Fig. 5D), A 3-D computer reconstruction (Imaris Bitplane) of an arterial segment identifies the eNOS-hemoglobin complexes to occupy regions of the endothelial cell that traverse the IEL (Fig. 5E); these regions are consistent with the size and location of myoendothelial junctions.

We next observed that each endothelial cell appeared to have a single subcellular domain containing the eNOS-hemoglobin complexes (Supplemental Fig. 4, A-D). The ratio of autofluorescent punctates to endothelial cell nuclei was consistent across artery segments from three different donors (1.21, 1.15, and 1.20), providing a mean ± SEM of 1.19 ± 0.03 (Supplemental Fig. 4 G).We then calculated the spatial density of eNOS-hemoglobin-containing punctates by counting the number of punctates per arterial surface area (Supplemental Fig. 4 E). The mean density was 0.0021 ± 0.0004 complexes/µm^2^ and was consistent across seven donors. To investigate whether fluorescent intensities of the eNOS-hemoglobin-containing punctates were consistent between two different arteries taken from the same donor, we imaged five arteries collected from three omental tissue donors. Arteries from the same donor did not differ in intensity, but one donor out of three had lower signal intensities than the other two (Supplemental Fig. 4 F).

**Figure 5.**
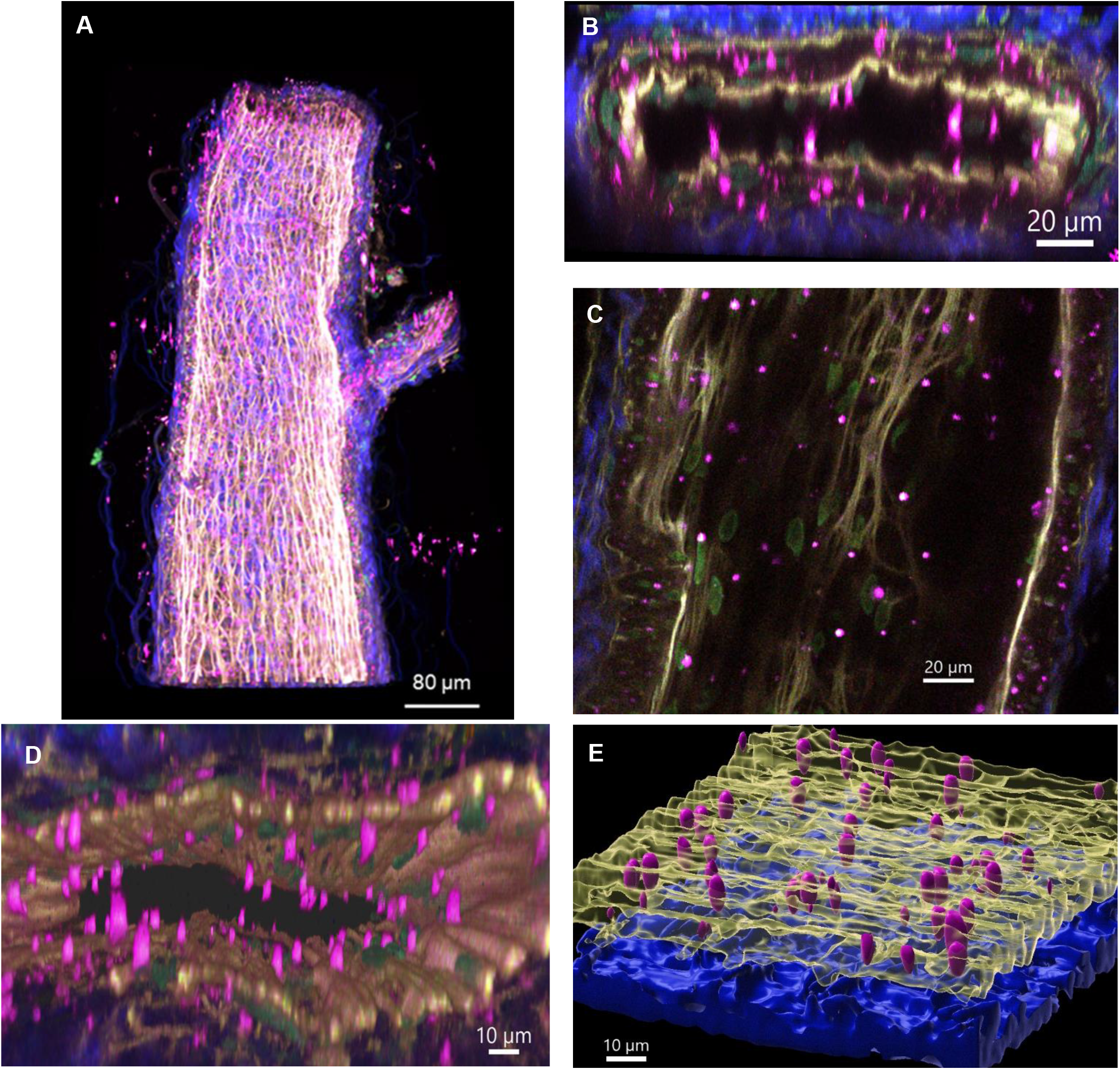
**(A-E):** Multiphoton imaging of intact omental arteries. **A)** Unlabeled intact omental artery. The collagen of the tunica externa demonstrates second harmonic generation signal (SHG) in the blue part of the spectrum; the elastin of the internal elastic lamina (IEL) us autofluorescent in the green and red spectra, showing here as yellow. **B)** Transverse cross section, 3µm in the Z-plane, of intact small omental artery shown in panel A, labeled with DAPI to demarcate nuclei (green). The transverse section shows localization of autofluorescent hemoglobin-eNOS complexes, which fluoresce broadly (shown here in pink) to the IEL (yellow). **C)** Longitudinal cross section, taken as 1 µm thick Z-step image of omental artery at the midline shows, the endothelial cell nuclei (green) aligned in parallel with the direction of the artery. The autofluorescent complexes are in the same 1µm plane as endothelial nuclei and the IEL (yellow). **D)** Three-dimensional image of segment from intact omental artery in panel A, viewed in the transverse plane and extending for 40µm. Endothelial nuclei are visible interior to the IEL, while smooth muscle nuclei are visible between the IEL and the collagen of the tunica externa. The autofluorescent complexes (pink) are localized to the IEL. **E)** Image deconvolution of omental artery, opened and imaged *en face*, as a three-dimensional surface model using Imaris Bitplane. Hemoglobin-eNOS complexes are visualized in pink, and are embedded within, and protruding through, the IEL (yellow). The collagen of the tunica externa is shown in blue.

### Disruption of the eNOS-hemoglobin complex enhances feedback vasodilation to an alpha-adrenergic agonist in human omental resistance arteries

We hypothesized that disruption of the hemoglobin/eNOS complex would acutely enhance feedback vasodilation from NO released from eNOS at the MEJ in response to alpha-1-adrenergic stimulation because the displacement of hemoglobin would render it less effective at scavenging NO produced by eNOS. To test this hypothesis, we performed *ex vivo* pressure myography on pairs of omental arteries obtained from each of five donors in response to escalating doses of the alpha adrenoreceptor agonist phenylephrine (responses fit with four parameter logistic regression best-fit values, Fig. 6; Supplemental Table 1). Initially, omental arteries in each pair maximally constricted to phenylephrine to a similar degree (4-PL minimum, 39.1 ± 3.2% vs 46.1 ± 5.5% of baseline diameter; p = 0.30; Fig. 6 A,B). Next, in one vessel from each donor pair, vasoconstriction was measured after incubation with the alpha globin mimetic peptide HbaX, which was previously established to disrupt binding between the alpha subunit of hemoglobin and eNOS^29,30^. The second vessel from each donor pair was treated with a control peptide before phenylephrine responses were measured. Arteries exposed to the alpha globin mimetic peptide maximally constricted less than those exposed to the control peptide (64.6 ± 1.6% vs 39.1 ± 3.2%; p < 0.01). To determine whether the enhanced feedback vasodilation was NOS-dependent, each artery was incubated with a combination of the peptide plus the NOS inhibitor L-NAME, and vasoconstriction to PE measured a third time. Following the incubation of mimetic peptide-treated arteries with L-NAME, phenylephrine-induced vasoconstriction was restored to 41.9 ± 2.0% of baseline (p < 0.0001 vs mimetic peptide alone), no different from the initial phenylephrine-induced response (p = 0.53). L-NAME had no effect on the arteries treated with the control peptide (p = 0.99). Thus, disruption of the hemoglobin-eNOS complex enhanced feedback vasodilation to alpha-1-adrenergic stimulation in a NOS-dependent manner.

**Figure 6:**
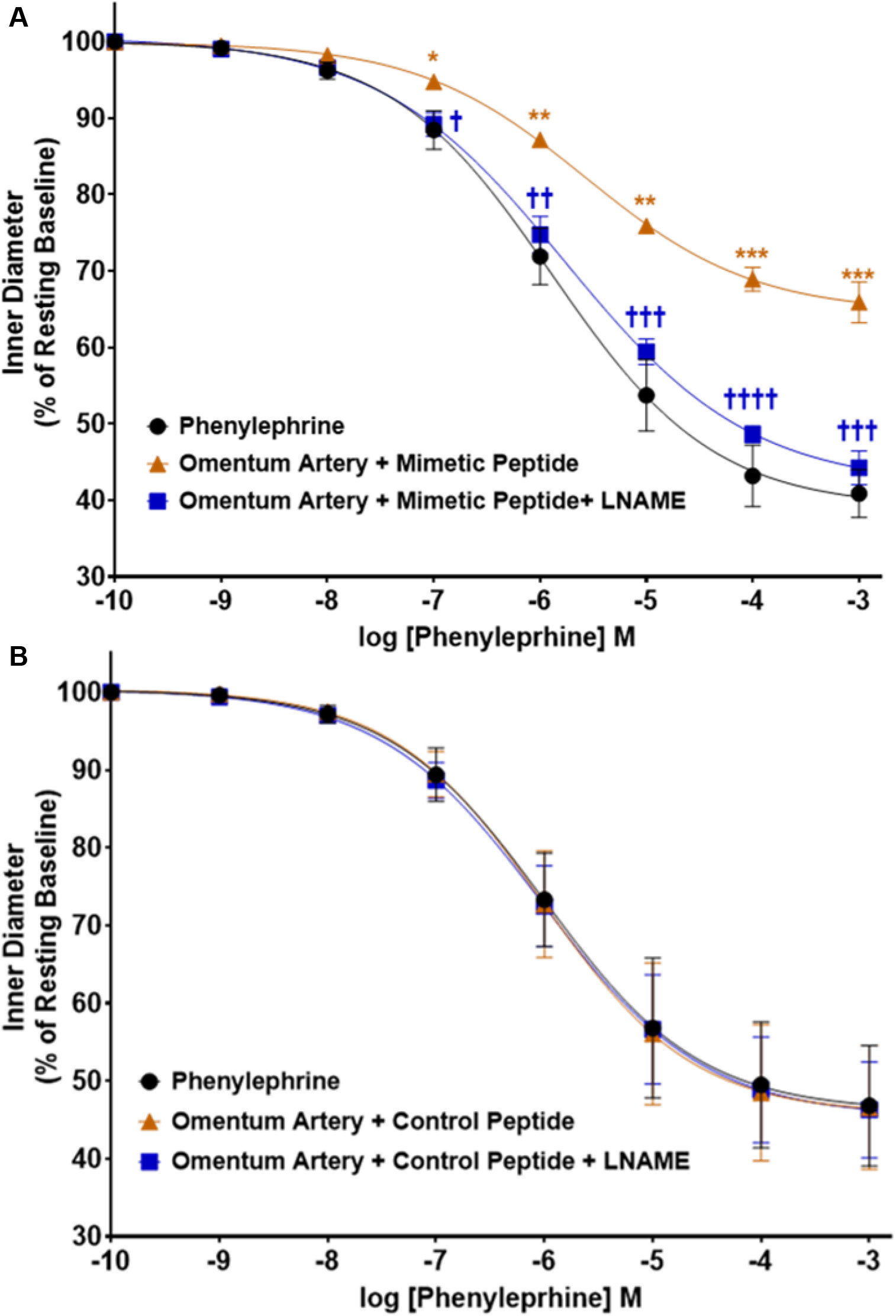
Change in inner diameter (as percentage of resting baseline) of omental arteries to phenylephrine (PE) with four-parameter logistic regression models fit to the data. Responses are plotted as (Mean Diameter, SE) Responses to only PE were not different between the two groups (p=0.66). **A)** Responses of omental arteries (n=5) to PE, PE after incubation with an alpha globin mimetic peptide, and PE after incubation with the mimetic peptide and L-NAME. Treatment with the mimetic peptide significantly reduced the PE response at baseline and in the presence of L-NAME (p<0.0001 for each). **B)** Responses of omental arteries (n=5) to PE, PE after incubation with a control peptide, and PE after incubation with the control peptide and L-NAME. There was not significant difference between response to PE following treatment with the control peptide (p=0.92) or the peptide and L-NAME (p=0.99). The response of the arteries at each dose of PE was then analyzed by 2-way Repeated Measures ANOVA. Significance between the peptide-only artery and initial PE response is denoted by green stars (* p<0.05; **p<0.01, ***p<0.001, ****p<0.0001); comparison between the L-NAME + peptide treated artery against the peptide-only artery is denoted by blue daggers († p<0.05; ††p<0.01, †††p<0.001, †††† p<0.0001)

## DISCUSSION

Blood pressure and blood flow are regulated by the dynamic and homeostatic behavior of resistance arteries. A major vasoconstrictive signal, alpha-1-adrenergic stimulation of vascular smooth muscle, is counterbalanced by the coupled release of vasodilatory signals such as nitric oxide (NO) from the adjacent endothelial cell.^18,21,25^ In 2012, Straub et al proposed a novel vasoregulatory mechanism whereby this coupled release of NO from endothelial cells is limited by monomeric alpha globin.^29^ That mechanism was based primarily on studies of cultured cells and murine arteries, and had yet to be systematically evaluated in the human arterial context. Given the potential importance of this regulatory mechanism to human circulatory physiology, we set out to understand the expression, binding partners, sub-cellular localization, and vasoregulatory function of alpha globin in human resistance arteries.

Human resistance arteries were dissected from fresh omental tissue, individually cannulated and perfused to remove blood, and then subjected to a range of molecular, biochemical, imaging, and functional studies. This systematic study, carried out entirely in the context of human resistance arteries, independently verifies the mechanism originally proposed by Straub et al.^29^ and extends it into the context of normal human arterial function. The data presented here suggest that hemoglobin, not alpha globin alone, regulates eNOS-dependent NO signaling between endothelium and smooth muscle of human resistance arteries.

Evidence supporting the discovery of hemoglobin in the human resistance artery wall came from multiple orthogonal approaches. First, we looked for endogenous expression of the alpha globin (*HBA1, HBA2*) and beta globin (*HBB*) genes in blood-free arterial tissue, and found all three genes to be expressed at a level similar to endothelial nitric oxide synthase (*NOS3*), a major endothelial gene. This result is in contrast to a study of thoracodorsal arteries from mice that detected *HBA* but not *HBB* transcripts,^31^ and there have been no studies of globin gene expression within perfused human arteries for comparison. To exclude the possibility that these alpha and beta globin transcripts originated from residual blood cells in the perfused arteries, we examined Band3 (*SLC4A1*), a gene expressed predominantly in red cell progenitors. *SLC4A1* transcripts were very low or absent from each arterial specimen; moreover, the ratio of *HBA1*:*SLC4A1* was much higher in arterial tissue than in whole blood, indicating that the *HBA1* transcripts could not have originated from blood contamination. Interestingly, the ratio of *HBA1*:*HBA2* was also greater in arterial tissue compared to blood, suggesting that the alpha globin genes are under endothelial cell-specific transcriptional control that differs from that of red blood cell progenitors. Thus we concluded that both alpha and beta globin genes are expressed in cells that make up the wall of human resistance arteries.

Next, we sought to understand what proteins interact with alpha globin in the context of its role in the arterial wall. In order to obtain sufficient amounts of protein to analyze by Western blot, we individually cannulated, perfused, and lysed more than 200 arteries from multiple donors and pooled them for protein analysis. Both eNOS and beta globin co-immunoprecipitated with alpha globin, implying that these three proteins form a stable complex *in vivo*. This result is in contrast to prior studies in mouse arteries where eNOS, but not beta globin, co-immunoprecipitated with alpha globin.^29^ Our results are also in contrast to a study that examined a small number of human subcutaneous adipose arteries by Western blot without co-immunoprecipitation and concluded that alpha globin, but not beta globin, was present.^32^ A third study detected more beta globin than alpha globin on a Western blot of protein extracted from human pulmonary arteries, but beta globin was not recognized to be of vascular origin.^43^

To further examine the biophysical interactions between alpha globin and eNOS, we used biolayer interferometry and found them to interact with moderate affinity in solution. The equilibrium dissociation constant of full-length alpha globin and the eNOS oxygenase domain had not previously been quantified, but it was similar to that previously reported for an alpha globin-derived peptide and eNOS oxygenase domain.^32^ Further evidence for the interaction of alpha globin with eNOS *in vivo* was provided by the observation of FRET between fluorophore-labeled antibodies against alpha globin and eNOS *in situ* in the arterial wall. Together, these three orthogonal approaches support the novel observation that alpha globin, beta globin, and eNOS interact and exist as a multi-protein complex in the human omental artery wall.

We also detected Cyb5R3, an enzyme that regulates the oxidative state of hemoglobin, to co-immunoprecipitate with alpha globin in protein lysates from perfused human resistance arteries. Cyb5R3 reduces the heme iron from the ferric (Fe^3+^) state to the ferrous (Fe^2+^) state which has a higher affinity for NO; thus Cyb5R3 could be an important redox regulator of nitric oxide signaling by the hemoglobin-eNOS complex.^29,44^

Molecular modeling of eNOS, alpha globin, beta globin and Cyb5R3 accommodates two heterotetrameric hemoglobin molecules bound to an eNOS oxygenase homodimer. The predicted interface between alpha globin and eNOS shifts relative to the model that incorporated an alpha globin monomer alone (Supplemental Fig. 5A).^29,30^ The revised model identifies novel interfaces between beta globin and eNOS, as well as between beta subunits from different hemoglobin tetramers (Supplemental Fig. 6A-C). The Cyb5R3 FAD binding domain can be brought in close proximity to both the alpha globin heme and eNOS reaction center to potentially facilitate the redox regulation of the alpha globin heme and its ability to scavenge nitric oxide (Supplemental Fig. 6E).

To understand the sub-cellular localization of this complex within the artery wall, we employed multiphoton (MP) microscopy. The primary advantage of this technique was that it enabled high-resolution imaging of intact arteries without sectioning; a secondary advantage was the ability of MP excitation to highlight structural features of the artery wall such as collagen (via SHG signal) and elastin (via narrow band autofluorescence). A third and unanticipated feature of MP microscopy was the excitation of hemoglobin to produce a broad-spectrum autofluorescence. We observed distinct, small punctates of intense autofluorescence in the wall of human arteries consistent with the emissions from tetrameric hemoglobin, but the same technique did not reveal autofluorescent punctates in mouse mesenteric arteries, agreeing with multiple prior observations that the beta chain of hemoglobin is absent in mouse arteries.^29,31^ In a non-perfused artery, these punctates were distinguishable from red blood cells by their size, intensity, and location. We verified that this autofluorescent complex contained alpha globin, beta globin, and eNOS by measuring the fluorescence lifetimes of photons emitted from fluorophore-labeled antibodies that were distinct from the lifetimes of photons emitted from autofluorescent hemoglobin. This hemoglobin-eNOS complex localized to a small region of each endothelial cell that penetrated the internal elastic lamina, a location that is consistent with the anatomical structure of the myoendothelial junction. This localization is in agreement with previous studies that identified an eNOS-alpha globin complex at the myoendothelial junction in co-cultured vascular cells and in the mouse thoracodorsal artery wall.^29^ Intact human resistance arteries have not previously been imaged for alpha globin or beta globin for comparison; however, a prior study that employed immunohistochemical analysis of mesenteric artery sections on a human tissue microarray did not detect beta globin.^31^ The localization of hemoglobin-eNOS to the myoendothelial junction of intact arteries fits with hemoglobin’s emerging role as a regulator of directed nitric oxide signaling between endothelium and smooth muscle in human resistance arteries.

We hypothesized that the contribution of NO during feedback vasodilation, whereby MEJ-localized eNOS produces NO to counteract vasoconstriction, would be regulated by the hemoglobin bound to eNOS. To stimulate eNOS at the MEJ, we used an alpha-1-adrenergic agonist, phenylephrine, known to activate eNOS at the MEJ via calcium and/or IP_3_ signals conducted from vascular smooth muscle.^18,21,25^Using isolated vessel pressure myography, we observed that human omental arteries constricted consistently to phenylephrine; however, treatment with an alpha globin mimetic peptide previously established to disrupt binding between eNOS and alpha globin^30,32^ diminished this constriction. This is consistent with an observation made by Keller et al, on four subcutaneous adipose arterioles obtained from hypertensive donors in which the alpha globin mimetic peptide HbaX reduced the constrictive effect of phenylephrine.^32^ In the final phase of our myography experiments, we applied the alpha globin mimetic peptide HbaX in combination with a NOS inhibitor and determined that the ability of the mimetic peptide to enhance feedback vasodilation was dependent on the enzymatic activity of NOS. This novel experiment provided further evidence that hemoglobin regulates the diffusion of NO produced directly from eNOS in response to alpha-1-adrenergic vasoconstriction in human resistance arteries.

Together, these observations reinforce several key elements of the concept first proposed by Straub et al that endothelial alpha globin regulates nitric oxide signaling by binding to eNOS at the MEJ.^29,30^ In human arteries, we find hemoglobin bound to eNOS in a complex at the myoendothelial junction where it limits the diffusion of nitric oxide produced enzymatically by eNOS. Studies conducted with the HbaX alpha globin mimetic peptide reveal it to be a potent disruptor of NO-scavenging in human vessels, implying that binding between alpha globin and eNOS is pivotal even when beta globin is present. Nevertheless, we speculate that hemoglobin may interact with eNOS in ways that are different than alpha globin alone. In addition to potentially increasing the molecular stability of the complex, beta globin can stabilize the inherently unstable alpha globin chain potentially preventing aggregation of alpha globin monomers; hemoglobin may have an enhanced ability to catalyze the reaction between NO and molecular oxygen because of the additional heme groups in the beta chains; hemoglobin might be more readily reduced by cytochrome B5 reductase, a step necessary to maintain fast reaction rates between NO and molecular oxygen catalyzed by ferrous heme iron; and beta globin could potentially coordinate the binding of alpha globin with eNOS as predicted by modeling and even alter that binding via cooperative changes that could be oxygen-dependent. These potential implications of eNOS binding to tetrameric hemoglobin instead of monomeric alpha globin merit further investigation.

This investigation into the role of hemoglobin as a regulator of nitric oxide signaling across the myoendothelial junction of human omental arteries has strengths and limitations. A major constraint of studying arteries from live human donors was the limited availability of omental tissue. For example, in the gene expression studies, while we excluded circulating blood cells as the source of globin transcripts, but we did not carry out single cell analyses to determine which cell types in the artery wall express *HBA1, HBA2*, or *HBB*. Although we have identified both alpha and beta globin proteins as constituents of the complex with eNOS, and the autofluorescence observed is characteristic of tetrameric hemoglobin, we did not determine the stoichiometry of this complex and cannot rule out other constituents. Despite these limitations, the data presented here identify hemoglobin as a localized regulator of directed nitric oxide signaling across the myoendothelial junction in human resistance arteries.

In humans, the discovery of vascular hemoglobin raises the possibility that genetic polymorphism in not only the alpha globin genes but also the beta globin gene could potentially affect nitric oxide signaling and vascular function. This is especially interesting because while polymorphism sin the alpha globin genes are primarily deletions that reduce the level of expression (e.g., the spectrum of alpha thalassemia syndromes),^45^ polymorphisms in the beta globin gene not only affect level of expression (e.g., the beta thalassemia syndromes) but also affect the structure of the beta chain (e.g., HbS, HbC, and HbE),^46,47^ raising the possibility that these heritable differences that cause anemia by altering the expression or structure of globins in the red blood cell might also change vascular function by altering the binding of hemoglobin to eNOS, resulting in new vascular phenotypes that are just beginning to be recognized. For example, a recent study found that healthy individuals with alpha globin gene deletions have increased nitric oxide-mediated vascular perfusion.^48^ We recently discovered alpha globin gene deletions to be associated with protection from chronic kidney disease and end-stage kidney disease.^49^ These associations were independent of sickle cell trait and hemoglobin level, raising the possibility that protection is conferred via mechanisms involving the vascular functions of alpha globin. Loss of alpha globin expression could enhance endothelial nitric oxide signaling and counteract vasoconstriction from adrenergic signaling – pathways previously implicated in the pathophysiology of kidney disease. Further study of arteries from people with polymorphism in the alpha globin, beta globin, or *CYB5R3* genes could further elucidate the role of hemoglobin in regulating vascular function and in determining vascular disease risk.

## CONCLUSION

In summary, hemoglobin forms a complex with eNOS at myoendothelial junctions in human omental resistance arteries where it limits the diffusion of NO produced by eNOS in response to vasoconstriction by an alpha-1-adrenergic stimulus.

## Supporting information

Full Methods and Supplemental Data

## Data Availability

Data are available from the authors by request

## ACKNOWLEDGEMENTS

We would like to thank Dr. Owen Schwartz, Head of the Biological Imaging Facility of the Research Technologies Branch, National Institute of Allergy and Infectious Diseases (NIAID-RTB), for his expertise and assistance with performing the multiphoton and Fluorescence Lifetime Imaging microscopy experiments.

The authors would like to thank the staff of the National Institutes of Health Clinical Center (NIH-CC) General Surgery Consultative Services for their assistance in coordinating tissue collection for this study.

The purified recombinant eNOS oxidase domain and purified alpha hemoglobin stabilizing protein used in the biolayer interferometry experiment was graciously provided by Christophe Lechauve, PhD, and Mitchell Weiss, MD PhD, of St. Jude Children’s Research Hospital, Hematology Department. The biolayer interferometry experiments were conducted with equipment and staff assistance from the National Heart, Lung, and Blood Institute Biochemistry Core.

Molecular graphics images were produced using the UCSF Chimera package from the Resource for Biocomputing, Visualization, and Informatics at the University of California, San Francisco (supported by NIH P41 RR-01081).

## Ethics Statement

The collection of human tissue specimens during clinically indicated abdominal operations, and use in this study, was approved by the Institutional Review Board (National Institutes of Health, National Cancer Institute) on IRB protocol 13-C-0176 registered at https://www.clinicaltrials.gov with unique identifier NCT01915225.

## References

1. Mulvany MJ, Aalkjaer C. Structure and function of small arteries. Physiol Rev. 1990;70:921–961.

2. Harper D, Chandler B. Splanchnic circulation. BJA Educ. 2016;16:66–71.

3. Thomas GD. Neural control of the circulation. Adv Physiol Educ. 2011;35:28–32.

4. Volianitis S, Secher NH. Cardiovascular control during whole body exercise. J Appl Physiol. 2016;121:376–390.

5. Hilgers RHP, Mey JD. Myoendothelial coupling in the mesenteric arterial bed; segmental differences and interplay between nitric oxide and endothelin-1. Br J Pharmacol. 2009;156:1239–1247.

6. Sun D, Messina EJ, Kaley G, Koller A. Characteristics and origin of myogenic response in isolated mesenteric arterioles. Am J Physiol-Heart Circ Physiol. 1992;263:H1486–H1491.

7. Shimokawa H, Yasutake H, Fujii K, Owada MK, Nakaike R, Fukumoto Y, Takayanagi T, Nagao T, Egashira K, Fujishima M, Takeshita A. The Importance of the Hyperpolarizing Mechanism Increases as the Vessel Size Decreases in Endothelium-Dependent Relaxations in Rat Mesenteric Circulation. J Cardiovasc Pharmacol. 1996;28:703–711.

8. Seals DR, Taylor JA, Ng AV, Esler MD. Exercise and aging: autonomic control of the circulation. Med Sci Sports Exerc. 1994;26:568–576.

9. Kregel KC. Augmented mesenteric and renal vasoconstriction during exercise in senescent Fischer 344 rats. J Appl Physiol. 1995;79:706–712.

10. Furchgott RF, Vanhoutte PM. Endothelium-derived relaxing and contracting factors. FASEB J. 1989;3:2007–2018.

11. Nava E, Llorens S. The paracrine control of vascular motion. A historical perspective. Pharmacol Res. 2016;113:125–145.

12. Segal SS. Regulation of Blood Flow in the Microcirculation. Microcirculation. 2005;12:33–45.

13. Reid JL. Alpha-adrenergic receptors and blood pressure control. Am J Cardiol. 1986;57:E6–E12.

14. Exton JH. Mechanisms involved in alpha-adrenergic phenomena. Am J Physiol-Endocrinol Metab. 1985;248:E633–E647.

15. Minneman KP. Alpha 1-adrenergic receptor subtypes, inositol phosphates, and sources of cell Ca2+. Pharmacol Rev. 1988;40:87–119.

16. Davis MJ, Hill MA. Signaling Mechanisms Underlying the Vascular Myogenic Response. Physiol Rev. 1999;79:387–423.

17. Wu D, Katz A, Lee CH, Simon MI. Activation of phospholipase C by alpha 1-adrenergic receptors is mediated by the alpha subunits of Gq family. J Biol Chem. 1992;267:25798–25802.

18. Dora KA, Doyle MP, Duling BR. Elevation of intracellular calcium in smooth muscle causes endothelial cell generation of NO in arterioles. Proc Natl Acad Sci U S A. 1997;94:6529–6534.

19. Dora KA, Hinton JM, Walker SD, Garland CJ. An indirect influence of phenylephrine on the release of endothelium-derived vasodilators in rat small mesenteric artery. Br J Pharmacol. 2000;129:381–387.

20. Nelson MT, Patlak JB, Worley JF, Standen NB. Calcium channels, potassium channels, and voltage dependence of arterial smooth muscle tone. Am J Physiol-Cell Physiol. 1990;259:C3–C18.

21. Garland CJ, Bagher P, Powell C, Ye X, Lemmey HAL, Borysova L, Dora KA. Voltage-dependent Ca2+ entry into smooth muscle during contraction promotes endothelium-mediated feedback vasodilation in arterioles. Sci Signal. 2017;10. doi:10.1126/scisignal.aal3806.

22. Lamboley M, Pittet P, Koenigsberger M, Sauser R, Bény J-L, Meister J-J. Evidence for signaling via gap junctions from smooth muscle to endothelial cells in rat mesenteric arteries: possible implication of a second messenger. Cell Calcium. 2005;37:311–320.

23. Isakson BE, Ramos SI, Duling BR. Ca2+ and inositol 1,4,5-trisphosphate-mediated signaling across the myoendothelial junction. Circ Res. 2007;100:246–254.

24. Yashiro Yasuaki, Duling Brian R. Integrated Ca2+ Signaling Between Smooth Muscle and Endothelium of Resistance Vessels. Circ Res. 2000;87:1048–1054.

25. Hong Kwangseok, Cope Eric L., DeLalio Leon J., Marziano Corina, Isakson Brant E., Sonkusare Swapnil K. TRPV4 (Transient Receptor Potential Vanilloid 4) Channel–Dependent Negative Feedback Mechanism Regulates Gq Protein–Coupled Receptor–Induced Vasoconstriction. Arterioscler Thromb Vasc Biol. 2018;38:542–554.

26. Tran CHT, Taylor MS, Plane F, Nagaraja S, Tsoukias NM, Solodushko V, Vigmond EJ, Furstenhaupt T, Brigdan M, Welsh DG. Endothelial Ca2+ wavelets and the induction of myoendothelial feedback. Am J Physiol-Cell Physiol. 2012;302:C1226–C1242.

27. Looft-Wilson RC, Todd SE, Araj CA, Mutchler SM, Goodell CAR. Alpha1-adrenergic-mediated eNOS phosphorylation in intact arteries. Vascul Pharmacol. 2013;58:112–117.

28. Boric MP, Figueroa XF, Donoso MV, Paredes A, Poblete I, Huidobro-Toro JP. Rise in endothelium-derived NO after stimulation of rat perivascular sympathetic mesenteric nerves. Am J Physiol-Heart Circ Physiol. 1999;277:H1027–H1035.

29. Straub AC, Lohman AW, Billaud M, Johnstone SR, Dwyer ST, Lee MY, Bortz PS, Best AK, Columbus L, Gaston B, Isakson BE. Endothelial cell expression of haemoglobin α regulates nitric oxide signalling. Nature. 2012;491:473–477.

30. Straub Adam C., Butcher Joshua T., Billaud Marie, Mutchler Stephanie M., Artamonov Mykhaylo V., Nguyen Anh T., Johnson Tyler, Best Angela K., Miller Megan P., Palmer Lisa A., Columbus Linda, Somlyo Avril V., Le Thu H., Isakson Brant E. Hemoglobin α/eNOS Coupling at Myoendothelial Junctions Is Required for Nitric Oxide Scavenging During Vasoconstriction. Arterioscler Thromb Vasc Biol. 2014;34:2594–2600.

31. Lechauve C, Butcher JT, Freiwan A, Biwer LA, Keith JM, Good ME, Ackerman H, Tillman HS, Kiger L, Isakson BE, Weiss MJ. Endothelial cell α-globin and its molecular chaperone α- hemoglobin-stabilizing protein regulate arteriolar contractility. J Clin Invest. 2018;128:5073–5082.

32. Keller TCS, Butcher JT, Broseghini-Filho GB, Marziano C, DeLalio LJ, Rogers S, Ning B, Martin JN, Chechova S, Cabot M, Shu X, Best AK, Good ME, Simão Padilha A, Purdy M, Yeager M, Peirce SM, Hu S, Doctor A, Barrett E, Le TH, Columbus L, Isakson BE. Modulating Vascular Hemodynamics With an Alpha Globin Mimetic Peptide (HbαX). Hypertens Dallas Tex 1979. 2016. doi:10.1161/HYPERTENSIONAHA.116.08171.

33. Sanner MF, Olson AJ, Spehner J-C. Reduced surface: An efficient way to compute molecular surfaces. Biopolymers. 1996;38:305–320.

34. Pettersen EF, Goddard TD, Huang CC, Meng EC, Couch GS, Croll TI, Morris JH, Ferrin TE. UCSF ChimeraX: Structure visualization for researchers, educators, and developers. Protein Sci. 2021;30:70–82.

35. van Zundert GCP, Rodrigues JPGLM, Trellet M, Schmitz C, Kastritis PL, Karaca E, Melquiond ASJ, van Dijk M, de Vries SJ, Bonvin AMJJ. The HADDOCK2.2 Web Server: User-Friendly Integrative Modeling of Biomolecular Complexes. J Mol Biol. 2016;428:720–725.

36. Shannon JP, Kamenyeva O, Reynoso GV, Hickman HD. Intravital Imaging of Vaccinia Virus-Infected Mice. In: Mercer J, ed. Vaccinia Virus: Methods and Protocols. New York, NY: Springer; 2019: 301–311.

37. Shahid M, Buys ES. Assessing Murine Resistance Artery Function Using Pressure Myography. JoVE J Vis Exp. 2013;:e50328.

38. Butcher JT, Goodwill AG, Frisbee JC. The ex vivo Isolated Skeletal Microvessel Preparation for Investigation of Vascular Reactivity. JoVE J Vis Exp. 2012;:e3674.

39. Jadeja RN, Rachakonda V, Bagi Z, Khurana S. Assessing Myogenic Response and Vasoactivity In Resistance Mesenteric Arteries Using Pressure Myography. JoVE J Vis Exp. 2015;:e50997.

40. Bryk AH, Wiśniewski JR. Quantitative Analysis of Human Red Blood Cell Proteome. J Proteome Res. 2017;16:2752–2761.

41. Rahaman MdM, Reinders FG, Koes D, Nguyen AT, Mutchler SM, Sparacino-Watkins C, Alvarez RA, Miller MP, Cheng D, Chen BB, Jackson EK, Camacho CJ, Straub AC. Structure Guided Chemical Modifications of Propylthiouracil Reveal Novel Small Molecule Inhibitors of Cytochrome b5 Reductase 3 That Increase Nitric Oxide Bioavailability*. J Biol Chem. 2015;290:16861–16872.

42. Zheng W, Li D, Zeng Y, Luo Y, Qu JY. Two-photon excited hemoglobin fluorescence. Biomed Opt Express. 2011;2:71–79.

43. Alvarez RA, Miller MP, Hahn SA, Galley JC, Bauer E, Bachman T, Hu J, Sembrat J, Goncharov D, Mora AL, Rojas M, Goncharova E, Straub AC. Targeting Pulmonary Endothelial Hemoglobin α Improves Nitric Oxide Signaling and Reverses Pulmonary Artery Endothelial Dysfunction. Am J Respir Cell Mol Biol. 2017;57:733–744.

44. Gladwin MT, Kim-Shapiro DB. Vascular biology: Nitric oxide caught in traffic. Nature. 2012;491:344–345.

45. Ribeiro DM, Sonati MF. Regulation of human alpha-globin gene expression and alpha-thalassemia. Genet Mol Res GMR. 2008;7:1045–1053.

46. Origa R. β-Thalassemia. Genet Med. 2017;19:609–619.

47. Weatherall DJ, Williams TN, Allen SJ, O’Donnell A. The Population Genetics and Dynamics of the Thalassemias. Hematol Oncol Clin North Am. 2010;24:1021–1031.

48. Denton CC, Shah P, Suriany S, Liu H, Thuptimdang W, Sunwoo J, Chalacheva P, Veluswamy S, Kato R, Wood JC, Detterich JA, Khoo MCK, Coates TD. Loss of alpha-globin genes in human subjects is associated with improved nitric oxide-mediated vascular perfusion. Am J Hematol. 2020. doi:10.1002/ajh.26058.

49. Ruhl AP, Jeffries N, Yang Y, Naik RP, Patki A, Pecker LH, Mott B, Zakai NA, Winkler C, Kopp J, Lange LA, Irvin MR, Gutierrez OM, Cushman M, Ackerman HC. HBA Copy Number and Kidney Disease Risk among Black Americans: a Longitudinal Cohort Study. medRxiv. 2021;:2021.04.01.21254397.

