## Supplementary material for "Hemoglobin Interacts with Endothelial Nitric Oxide Synthase to Regulate Vasodilation in Human Resistance Arteries": Full Methods and Supplemental Data

### Methods and Data Supplement

**Appendix A: Full Materials and methods**

**Appendix B: Supplemental Data Figures and Tables**

**Appendix C: Multiphoton Microscopy Image Acquisition Settings**

### Appendix A: Full Materials and Methods

***Collection of omental arteries and subcutaneous adipose arteries:*** Human omental tissue was collected from patients during clinically indicated abdominal procedures at the NIH Clinical Center. All patients provided informed consent for tissue procurement on IRB-approved protocol 13-C-0176 (NCT01915225). In six patients, subcutaneous adipose tissue that was removed by the surgeon was also collected to enable gene expression comparison in an additional tissue. All tissue removed by the surgeon was immediately placed into cold Krebs-HEPES (KH) buffer. Tissue was submerged in cold 4 ˚C KH buffer and kept on ice during dissection and artery isolation. Small arteries were isolated and dissected away from tissue using fine tip forceps (Dumont #5, Fine Science Tools) under a binocular stereo microscope. Isolated arteries were cannulated on one end with a glass micropipette in an isolated microvessel perfusion chamber (Danish Myo Technologies) and gently perfused with cold KH buffer to remove RBCs from the vessel lumen. Arteries were then prepared for downstream application. For gene expression, arteries were placed into RNAlater and frozen at -80 ˚C. For Western blot, perfused arteries were flash frozen and stored at -80C. For multiphoton microscopy and FLIM, arteries were fixed in 4% formaldehyde solution in PBS and then processed for immunofluorescent labeling. For pressure myography, arteries were cannulated on both ends and warmed to 37˚C in KH buffer.

***Gene Expression*:** Gene expression was measured by reverse-transcriptase droplet digital PCR using ddPCR primer/probe assays for *HBA1*, *HBA2*, *HBB*, *NOS3*, and *SLC4A1*(Bio-Rad) in arteries from human omental tissue and human subcutaneous adipose tissue and human whole blood in PaxGene RNA tubes. Total mRNA was extracted from arteries frozen in RNAlater using Qiagen RNEasy Kits; arteries were thawed on ice and homogenized in lysis buffer at 2 ˚C (CryoLysis, Bertin Instruments). Blood stored in PaxGene RNA tubes was processed using the PaxGene mRNA extraction kit (PreAnalytix). Total mRNA from both arteries and whole blood was measured by Denovix. mRNA was converted to cDNA by SuperScript IV VILO (Invitrogen); 1ng of cDNA was loaded per droplet digital PCR reaction. A no-reverse transcriptase control reaction (No-RT) was performed as a control during cDNA conversion, and evaluated by ddPCR as a negative control. Gene expression was quantified as total transcripts of target gene per 1 ng of cDNA. Expression ratios were calculated from gene expression per 1 ng cDNA values; expression ratios between omental arteries and whole blood, and subcutaneous arteries and whole blood, were compared by Mann-Whitney test.

***Co-Immunoprecipitation and Western blot:*** Up to 30 freshly dissected, perfused small omental arteries were collected from each of nine individual donors and flash frozen. Fresh frozen human omentum arteries were grinded manually on the top of a liquid nitrogen bowl using a tissue grinder. Then grinded tissue powders were subjected to protein extraction using Minute™ Total Protein Extraction Kit for Blood Vessels (InventBiotech # SA-03-BV) according to the manufacturer's instructions. Protein concentration were determined by BCA assay (# 23227, ThermoFisher Scientific).

For western blotting, 20 µg of total blood vessels proteins were separated by SDS-PAGE on 4-12% gels. Proteins were transferred to PVDF membrane and blocked by 5% non-fat milk, then immunoblotted with anti-Band 3 polyclonal antibody (1:1000, Fisher Scientific #PA5-80030) at

4ºC overnight. The next day, the proteins were incubated with horseradish peroxidase (HRP)-conjugated secondary antibodies (Fisher Scientific) and detected by enhanced chemiluminescence (Pierce).

For studies of the protein and protein interaction of the alpha globin, beta globin, and eNOS complex, co-immunoprecipitation was performed. In brief, 600 µg of blood vessels lysates from two health volunteers were incubated with 4 µg alpha globin polyclonal antibody (HBA, 1:1000, Proteintech, #14537-1-AP) or normal rabbit IgG antibody (Cell signaling #2729) at 4ºC overnight. Magnet protein A/G beads washed three times with PBS buffer were added to the lysates at room temperature for 1 hour. The beads were pelleted by a magnet stand, followed by five washes with PBS buffer. Protein complex were eluted by lysis buffer with gentle rotation and SDS loading buffer was added. Sample were boiled for 5 min, and subsequently analyzed by SDS-PAGE and Western blotting with HBA antibody, beta globin polyclonal antibody (HBB, 1:1000, Proteintech # 16216-1-AP) and eNOS polyclonal antibody (1:1000, Cell Signaling #32027).

***Protein-protein binding affinity assessed by biolayer interferometry*:** Biolayer interferometry experiments were performed on the automated eight-channel Octet RED96 instrument (ForteBio). The temperature was set at 30 °C unless otherwise specified. Alpha globin was biotinylated using EZ-Link™ NHS-PEG4 Biotinylation Kit (Thermofisher Scientific). Specifically, alpha globin was diluted to 100 µg/mL in 500 µL of 1xPBS, then 2.37 µL of Biotin at 2 mM was added to the Alpha globin solution and incubated on ice for 2 hours. Biotinylated Alpha globin was purified using Zeba Spin desalting column with excess biotin removal. Further, Alpha globin and all other tested proteins in this assay were dialyzed against assay buffer (1 x PBS, pH 7.4, containing 0.1% BSA and 0.05% Tween 20) for 24 hours with three times of buffer change. The dialyzed protein was diluted to 10 µg/mL in assay buffer and immobilized onto Streptavidin (SA) Biosensors via the strong and highly specific interaction between the Biotin and Streptavidin. The loading of the ligand protein was within a linear range of BLI signal to protein concentrations. Shaking platforms were set at 1000. Sample volumes in all steps were provided in 200 µL aliquots for the 96-well formats. After loading, a baseline step was performed followed by association and dissociation of analytes and controls.

Binding kinetics and data traces were obtained using Data Analysis software v8.2 (ForteBio), after subtraction of reference sensors from the sensors exposed to analytes at different concentrations. The equation used to fit the association is based on that the rate of association is a function of the decreasing concentration of unbound ligand molecules as analyte binding occurs: $Y = Y_0 + A(1-e^{-k_{obs}*t})$, where Y= level of binding, $Y_0$ = binding at start of association, A is an asymptote and t is time. $k_{obs}$ is the observed rate constant, which reflects the overall rate of the combined association and dissociation of the two binding partners. The equation used to fit dissociation is $Y= Y_0 + Ae^{-k_d*t}$, where $Y_0$ is binding at start of dissociation, and $k_d$ is the dissociation rate constant, which is expressed in unites of $sec^{-1}$. The association rate constant, $k_a$, which represents the number of complexes formed per second in a 1 molar solution of ligand and analyte, is expressed in $M^{-1}sec^{-1}$. The equation for $k_a$ is: $k_a = (k_{obs} - k_d)/ [Analyte]$.

The affinity constant $K_D$ is calculated using $k_a$ and $k_d$: $K_D = k_d / k_a$. The $K_D$ corresponds to the concentration of analyte at which 50% of ligand binding sites are occupied at equilibrium and is expressed in molar units (M).

***Molecular modeling of alpha globin, hemoglobin, cytochrome B5 reductase, and eNOS:*** Molecular graphics and analyses were performed with UCSF Chimera, developed by the Resource for Biocomputing, Visualization, and Informatics at the University of California, San Francisco, with support from NIH P41-GM103311. Molecular surfaces were created with the MSMS package within Chimera.[37] Alignment of protein structures was performed using the MatchMaker tool in UCSF Chimera. Protein-protein docking was performed using the HADDOCK 2.4 online server: https://wenmr.science.uu.nl/haddock2.4.[38]

A model based on Figure 1 in Straub et al. (ATVB, 2014)[30] was created in order to identify general regions of contact between eNOS and alpha globin, particularly with the respect to the location of the HbaX alpha globin mimetic peptide. Computational docking of eNOS to alpha globin performed using HADDOCK was guided by specifying the residues from the HbaX peptide on alpha globin and the general area of eNOS that contacted alpha globin on the original model. The five best docking clusters by HADDOCK score were examined in more detail. An alpha chain from hemoglobin (PDB ID 6bb5) was aligned to the alpha globin chain from each of the HADDOCK models. The third best-scoring model did not have significant bad clashes with eNOS. Notably, this model also provided several regions of interaction between β-globin and eNOS; it was used in subsequent analysis.

The HADDOCK model was also aligned with the second eNOS subunit in eNOS/hemoglobin model to place a second alpha globin, and then an alpha chain from hemoglobin was aligned to this second alpha globin as described above to generate a complex with two hemoglobin proteins docked to an eNOS homodimer. While there are some clashes between the two hemoglobin monomers, in this arrangement there are large regions of surface contact between the beta globin subunits on the two hemoglobin structures. Some clashes between the beta globin monomers are not unexpected, since the docking only considered the alpha globin and eNOS crystal structures and any errors in the docked poses will be magnified with distance from the region of contact, e.g., in the region where the beta globin subunits interact.

A model based on Supplementary Figure 15 in Straub et al. (Nature, 2012)[29] was created in order to identify general regions of contact between Cyb5R3 (PDB Id 1UMK) and the eNOS/alpha globin docked model. The FAD reaction center points away from the heme binding sites on eNOS and alpha globin in this earlier low-resolution model. However, a putative model of Cyb5R3 with all 3 reaction centers accessible to a common central pocket was generated via manual modeling using the virtual reality mode of UCSF ChimeraX.[38] This type of modeling is facilitated in VR since it is straightforward to move one protein relative to others see all the areas of contact in a single view.

***Antibody labeling of intact omental arteries:*** Isolated and perfused small omental arteries were fixed in 4% formaldehyde in PBS solution (ImageIT, Invitrogen). Vessels were then washed in PBS. Vessels undergoing immunofluorescent labeling with one antibody were permeabilized with 0.1% Triton-X 100 and incubated overnight at 4˚C with primary conjugated antibodies for alpha globin (HBA-AlexaFluor 647, Abcam ab215919), beta globin (HBB-AlexaFluor 546 SantaCruz Biotechnology sc-21757-AF546), and endothelial nitric oxide synthase (eNOS-AlexaFluor 594, SantaCruz Biotechnology sc-376751-AF594). In select arteries, DAPI (Invitrogen) labeling was performed to demark cell nuclei. Vessels undergoing immunofluorescent labeling with multiple antibodies for measurement of fluorescence lifetime and possible fluorescent resonance were permeabilized with 0.1% Triton-X 100, blocked in goat serum, and incubated overnight at 4˚C with primary antibodies for alpha globin (rabbit monoclonal, Abcam ab92492), beta globin (mouse monoclonal, Santa Cruz Biotechnology sc-21757), and/or eNOS (mouse monoclonal, Abcam ab76198). Arteries were then washed and stained with secondary fluorescent antibodies for 1 hour at room temperature (AlexaFluor-488 anti-mouse, AlexaFluor-546 anti-rabbit, AlexaFluor-546 anti-mouse, or AlexaFluor-647 anti-rabbit).

***Multiphoton imaging of intact omental arteries*****:** Intact arteries were placed on cover-glass bottom microscope stage in a drop of carbomer-based solution to prevent drying and immobilized under a coverslip. Images were acquired using a Leica SP8 Inverted DIVE (Deep In Vivo Explorer) multiphoton (MP) microscope (Leica Microsystems, Buffalo Grove, IL) with 25.0X and 40.0X Water Immersion Objective as previously described.[33] Multiphoton excitation was performed at 880nm (MaiTai DeepSee, Spectra Physics) and 1150nm (InSight DeepSee, Spectra Physics) and emitted fluorescence was measured using a 4Tune 4 HyD (4 tunable non-descanned hybrid detectors (HyDs)) reflected light detector set for simultaneous detection at the following spectra: Channel 1 (393-472nm), set for Blue; Channel 2 (496-547nm), set for Green; Channel 3 (590-621nm), set for Red; Channel 4 (658-717nm), set for Far Red. Multiphoton images were collected as a Z-stack in 1-3µm steps for up to 50 single phases; for larger blood vessels, tiled images were collected using mosaic merge. The collected image files were exported into Huygens Pro SVI and Imaris Bitplane for image deconvolution, processing, and analysis.

***Fluorescence lifetime imaging microscopy (FLIM)*****:** Images were acquired using the Leica SP8 Inverted DIVE multiphoton microscope. FLIM images were collected using multiphoton excitation with Mai Tai-MP laser (Spectra Physics) tuned at 880nm at a 80Mhz frequency and images were simultaneously acquired for MP imaging and FLIM using 4-Tune External Hybrid detectors at wavelengths: Channel 1 (393-472nm); Channel 2 (496-547nm); Channel 3 (590-621nm); and Channel 4 (658-717nm). Arteries were labeled for eNOS, alpha globin, or eNOS, using antibodies labeled with AlexaFluor 488, AlexaFluor 568, or AlexaFluor 647. Images were acquired at 512-512-pixel format, collecting in excess of 2,000 photons per pixel. Fluorescent Decays and Förster resonance energy transfer (FRET/FLIM) efficiency transients and FRET-FLIM Images were collected, analyzed and processed using LASX Single Molecule detection analysis software. Fluorescence lifetime data for the entire collected image was plotted as histograms showing the totality of lifetime events recorded, with the peak lifetime and full-width, half-maximum (FWHM) reported for each histogram. Regions of interest were drawn around the globin-eNOS complexes using LASX Single Molecule detection analysis software, and mean lifetimes computed for each complex. Mean lifetimes of ROIs were compared by one-way AVOVA, with group means compared individually with post-hoc Tukey Test (GraphPad Prism v9.0).

***Multiphoton image processing and analysis of fluorescence signal intensity:*** The Leica Image File (.lif) for each artery was deconvolved to increase resolution and decrease noise and background (Huygens Pro, SVI). A region of interest was created manually by constructing a surface in the center of the artery that eliminated areas of tissue folding on sides  the outer edges (Imaris, Bitplane). This ROI was set to encompass the length of the artery in the Z plane (ADD Supplemental Image of Surfaces). Within the X and Y plane, the ROI included the internal elastic lamina, as well as the myoendothelial junctions of the vascular smooth muscle cells, and endothelial cell layers. The ROI excluded the vascular smooth muscle cell bodies and tunica externa. Within the ROI, the autofluorescent globin complexes were created as surface objects, based on fluorescence intensity in channels 2, 3, and 4. The globin complexes computed into automatic surface objects, with volumes defined by image voxels with positive fluorescent signal for each of these surface objects. Localization, volume, and mean fluorescence intensity statistics for each globin complex was calculated and exported from Imaris. Mean fluorescence intensity in each detector channel (Channel 1 (393-472nm), Channel 2 (496-547nm), Channel 3 (590-621nm), Channel 4 (658-717nm)) was plotted for each surface object, as well object size in voxels. The density of globin complexes was defined as the number of surface objects within the volume of the ROI surface. Mean intensity for each complex was plotted for each artery; as the mean intensity values were not normally distributed, densities for each artery were compared by Kruskal-Wallis with Dunn's multiple comparisons (GraphPad Prism v9.0). To determine the ratio of globin complexes to cell, arteries that had been stained by DAPI and labeled with HBA-AlexaFluor-647 were analyzed in Imaris, where Ch.2 and Ch. 3 were digitally subtracted, so that DAPI staining in Channel 1 and AlexaFluor-647 staining in Channel 4 remained. ROIs were drawn to encompass 20-30 endothelial cell nuclei (defined by shape and alignment with direction of flow in artery lumen) and exclude vascular smooth muscle cell nuclei (defined by shape and perpendicular alignment to luminal direction of flow). Three ROIs were drawn in each of three arteries, and the ratio of globin complexes to nuclei calculated for each ROI. Globin complex/endothelial cell nuclei ration was compared across the three arteries by one-way AVOVA, with group means compared individually with post-hoc Tukey Test (GraphPad Prism v9.0).

***Alpha globin mimetic peptide*: **A previously published, multi-species sequence homology analysis of alpha globin identified a conserved 10 amino acid sequence LSFPTTKTYF that was predicted to facilitate alpha globin binding with eNOS[30].  This sequence was combined with an N-terminal HIV-*tat* tag sequence (YGRKKRRQRRR) to facilitate membrane permeability, forming the alpha globin mimetic peptide (YGRKKRRQRRRLSFPTTKTYF, Anaspec). This peptide has been shown to disrupt association of alpha globin and eNOS *in vivo* in small arteries from mice[2], resulting in increased nitric oxide bioavailability and reduced constrictive potency of the alpha-1-adrenergic agonist phenylephrine in murine thoracodorsal arteries. A scrambled version of the peptide with an N-terminal HIV-*tat* tag sequence (YGRKKRRQRRRFPYFSTKLTT, Anaspec) was used as a peptide treatment control.

***Isolated artery pressure myography:*** For each myography experiment, two culture myograph wells (DMT-USA CM204) were prepared with 37°C Krebs-HEPES (KH) buffer. The pressure myography methodology utilized here for omental arteries is consistent with published protocols for assessing reactivity of mesenteric articles using a DMT pressure myograph[34]. Arteries 100-200µm in diameter were dissected from omental tissue immersed in cold KH buffer while on ice and transferred to culture myograph wells and warmed to 37°C. Arteries were cannulated with glass micropipettes and secured on both ends with 10-0 monofilament suture. Arteries were gradually pressurized to 60mmHg while perfused with KH buffer and allowed to equilibrate. The culture myograph chambers were then mounted on an inverted microscope at 10x magnification (DMT-USA). Digital video calipers were used to measure in the inner and outer diameter of cannulated arteries (MyoVIEW, DMT-USA).

Two arteries were used in parallel to enable simultaneous evaluation of the effect of the alpha globin mimetic peptide and control peptide with scrambled amino acid sequence. All arteries used developed at least 20% myogenic tone after pressurization, consistent with standards for pressure myography[34–36]. To test fitness of human arterial vascular smooth muscle for vasoconstriction, arteries were incubated with $10^{-4}$M KCl[38]; if the vessel did not constrict robustly to KCl, then the artery was discarded and replaced with another freshly dissected omental artery.

Arteries were then assessed for physiologic vasoconstriction response to the alpha-1-adrenoreceptor agonist phenylephrine (PE) (Sigma-Aldrich). Vasoconstriction was recorded to a PE dose response curve at effective concentrations of PE [$10^{-9}$, $10^{-8}$, $10^{-7}$, $10^{-6}$, $10^{-5}$, $10^{-4}$, $10^{-3}$] M. After PE washout, the arteries were allowed to re-equilibrate at 60mmHg. The arteries were then incubated in either alpha globin mimetic peptide (5 µmol/L in KH buffer) or the control peptide (5 µmol/L in KH buffer) for 45 minutes. The PE dose response curve was then repeated- PE [$10^{-9}$, $10^{-8}$, $10^{-7}$, $10^{-6}$, $10^{-5}$, $10^{-4}$, $10^{-3}$] M. Following washout and equilibration at 60mmHg, the two arteries were then incubated in the NOS inhibitor Nω-Nitro-L-arginine methyl ester HCl (L-NAME, $10^{-4}$ M)(Sigma-Aldrich) for 45 minutes. The PE dose response curve was then repeated for a final time- PE [$10^{-9}$, $10^{-8}$, $10^{-7}$, $10^{-6}$, $10^{-5}$, $10^{-4}$, $10^{-3}$] M.

The dose response of each artery was measured as baseline and post-incubation inner vessel lumen diameter. This response was normalized as percentage of baseline inner diameter. A sigmoidal four-parameter logistic regression, with X set on log base 10 scale, was fit to each dose response curve (GraphPad Prism v9.0). The four-parameter logistic models for each artery treatment condition were compared by extra sum-of-squares F test for each parameter in Prism to test the null hypothesis that each model and each parameter was significantly different between treatment conditions. The constrictive response between treatment conditions at each PE dose was also analyzed by two-way Repeated Measures ANOVA (GraphPad Prism v9.0). Significance was set for ($p<0.05$).

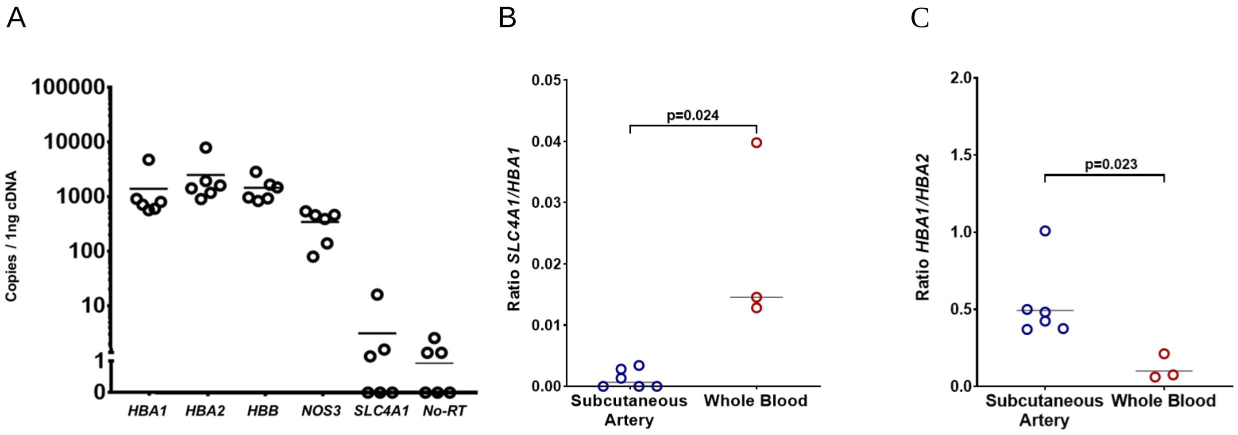


**Supplemental Figure 1:** Gene expression in perfused human small arteries dissected from subcutaneous adipose tissue. **A)** Total copies of *HBA1*, *HBA2*, *HBB, NOS3*, *SLC4A1* and a no-reverse transcriptase reaction control, per 1 ng of cDNA. **B)** The expression ratio of the erythrocyte specific marker *SLC4A1* to *HBA1* is lower in subcutaneous arteries (n=8) compared to human whole blood (n=3)(p=0.024), and **C)** the expression ratio *HBA1* to *HBA2* is lower in subcutaneous arteries (n=8) compared to human whole blood (n=3)(p=0.023).

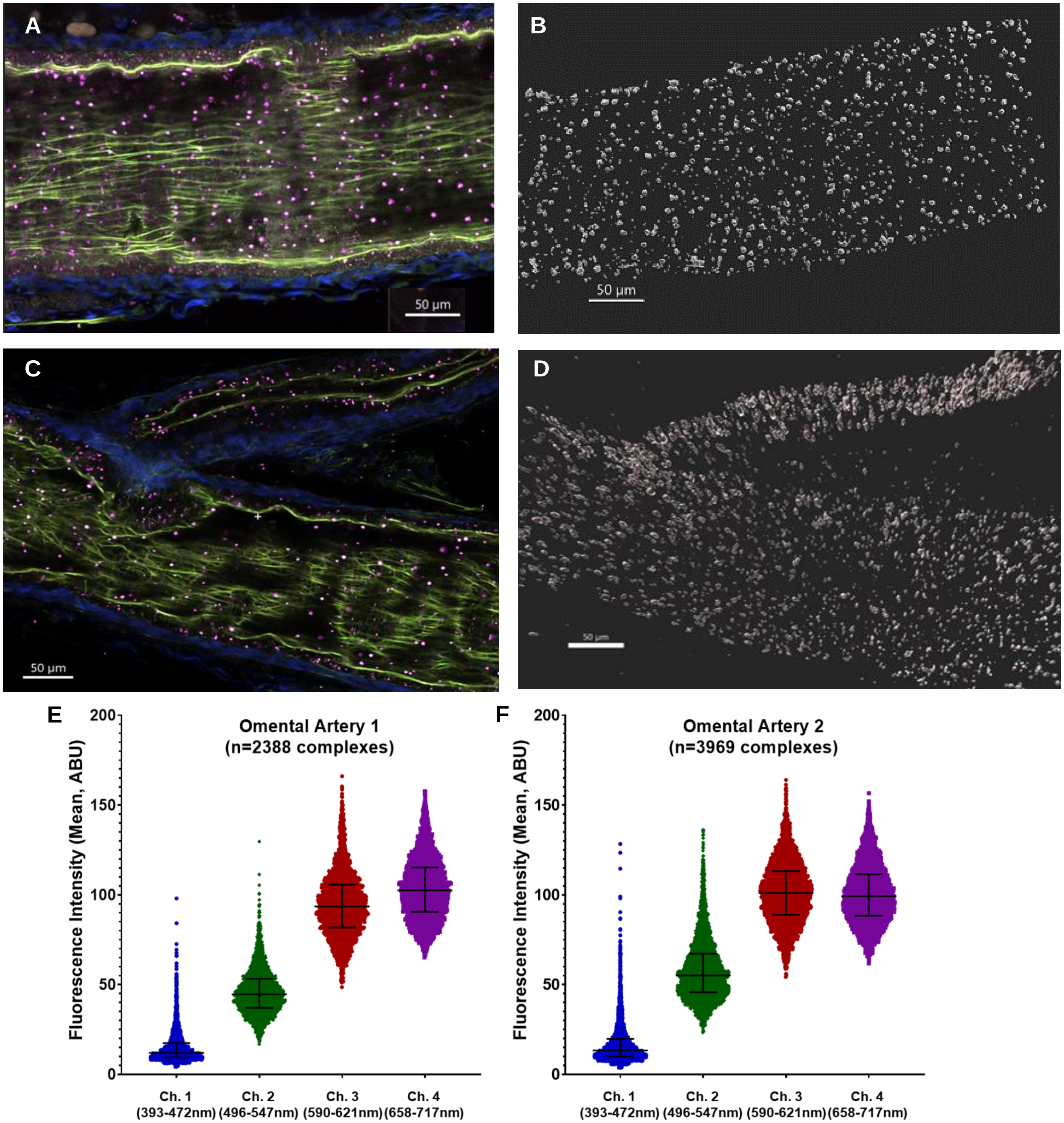


**Supplemental Figure 2: A)** Longitudinal view of unlabeled intact omental artery 1. Collagen is autofluorescence visible in blue; IEL as green; autofluorescent hemoglobin-eNOS complex is pink**. B)** Globin-eNOS complexes from entire intact omental artery 1 image are converted into individual surface objects in Imaris (n=2388). C) Longitudinal view of unlabeled intact omental artery 2, and D) Imaris conversion of globin-eNOS complexes to surface objects (n=3969). E) The mean fluorescence intensity in each channel is plotted for each surface object from omental artery 1; F) mean fluorescence intensity for surface objects from omental artery 2.

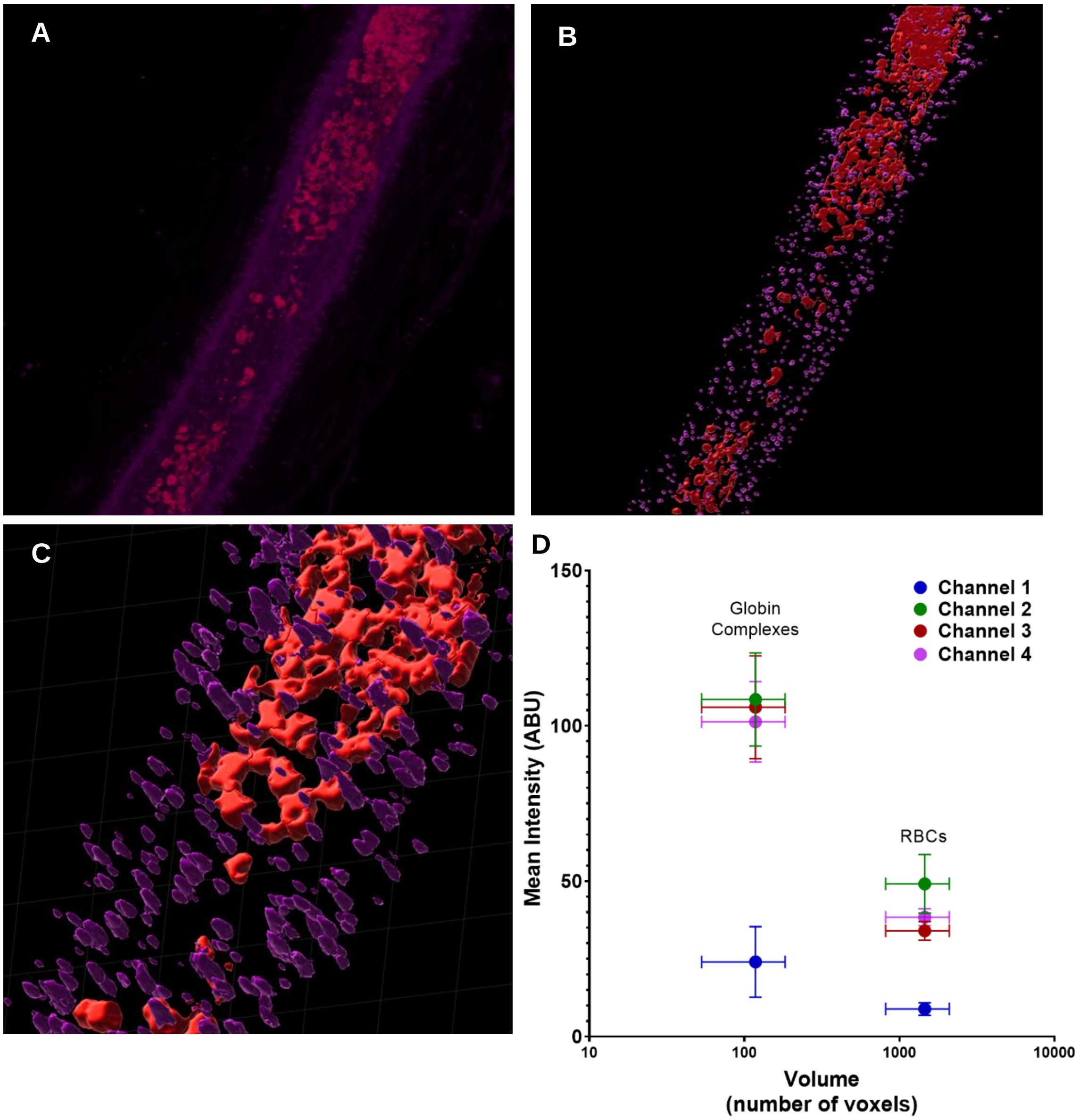


**Supplemental Figure 3 (A-D)**: Size and mean fluorescence intensity of hemoglobin-eNOS complexes and RBCs in the same intact omental artery. **A)** Artery image was enhanced in Imaris using size and spatial location filters to separate the RBCs from the hemoglobin-eNOS complexes. **B)** Distinct surface objects were created for each complex and each ROI. **C)** Zoomed view of surfaces shows delineating of objects encoded for RBCs vs hemoglobin-eNOS complexes. **D)** Mean fluorescence intensity plotted by mean volume for all hemoglobin-eNOS complexes compared to all RBCs.

E

F

G

H

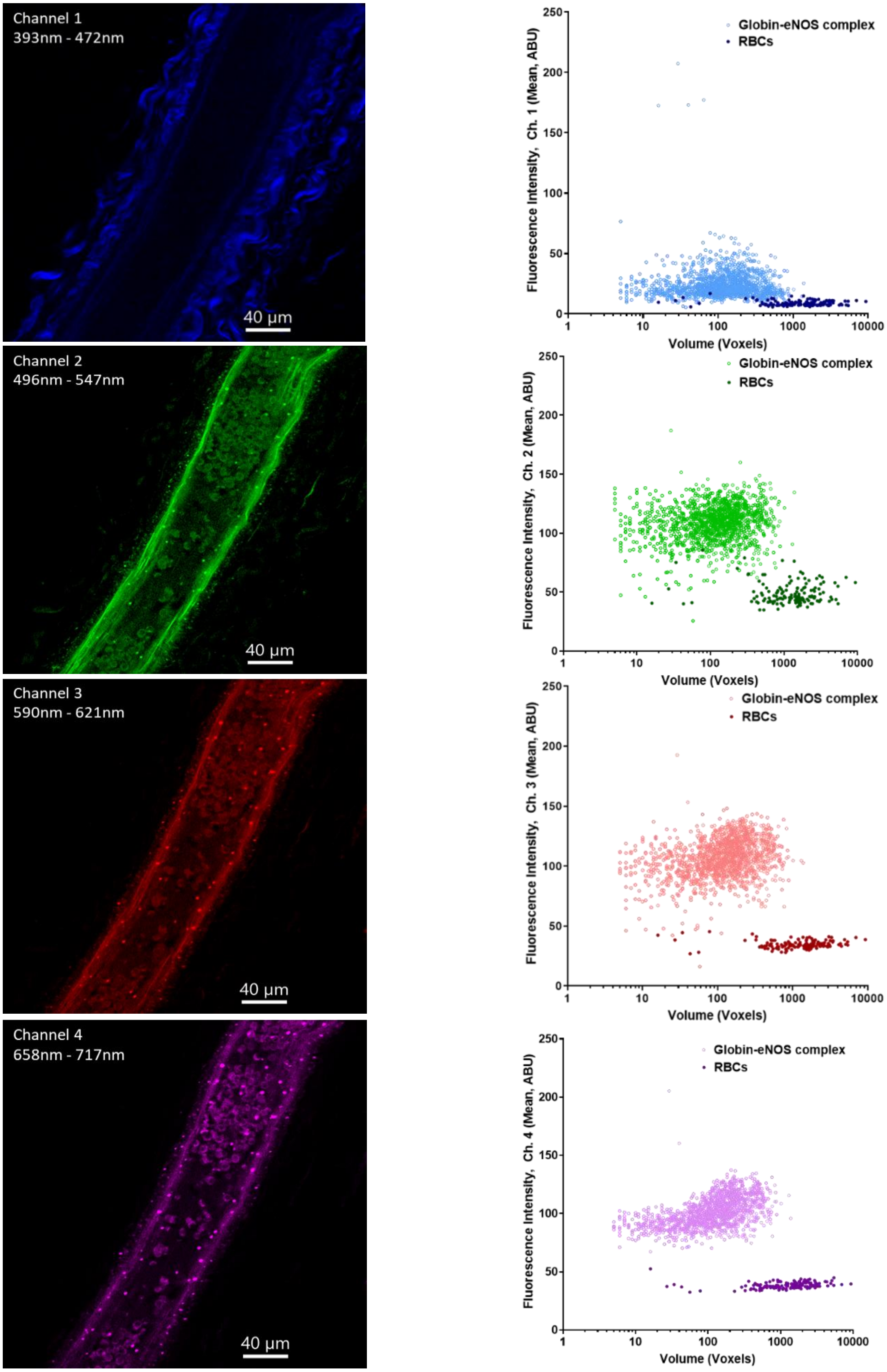

Channel 1
393nm - 472nm
40 µm
Fluorescence Intensity, Ch. 1 (Mean, ABU)
Globin-eNOS complex
RBCs
Volume (Voxels)
Channel 2
496nm - 547nm
40 µm
Fluorescence Intensity, Ch. 2 (Mean, ABU)
Globin-eNOS complex
RBCs
Volume (Voxels)
Channel 3
590nm - 621nm
40 µm
Fluorescence Intensity, Ch. 3 (Mean, ABU)
Globin-eNOS complex
RBCs
Volume (Voxels)
Channel 4
658nm - 717nm
40 µm
Fluorescence Intensity, Ch. 4 (Mean, ABU)
Globin-eNOS complex
RBCs
Volume (Voxels)

**Supplemental Figure 3 (E-H):** The autofluorescent hemoglobin-eNOS punctates have higher mean fluorescence intensity and smaller volume compared to RBCs within a non-perfused, intact omental artery.  Distinct surface objects were created as ROIs for each of the autofluorescent hemoglobin-eNOS complexes (n=2257) and separately the resident RBCs (n=142) using Imaris Bitplane (Supplemental Fig. 3, A-C). The autofluorescence of the punctates and RBCs within the range of each HyD detector are shown for the artery pictured in Figure 2B. **E)** Channel 1 detected autofluorescence from 393nm-472nm; **F)** Channel 2 detected autofluorescence from 496nm-547nm, **G)** Channel 3 detected autofluorescence from 590nm-621nm, **H)** Channel 4 detected autofluorescence from 658nm-717nm. The volume (voxels) and mean fluorescence intensity of each punctate and each RBC was calculated using Imaris (Bitplane) for each channel. The hemoglobin-eNOS complexes (median, 95%CI) [118 voxels, (113, 126)] were significantly smaller than the RBCs (p=0.0001) [1323 voxels, (1042-1538)]. Mean fluorescence intensity was lowest in Channel 1, but the intensities for the punctates and RBCs were clearly distinct in each channel, with punctates having higher mean intensities in all four channels (p=0.0001 for each).

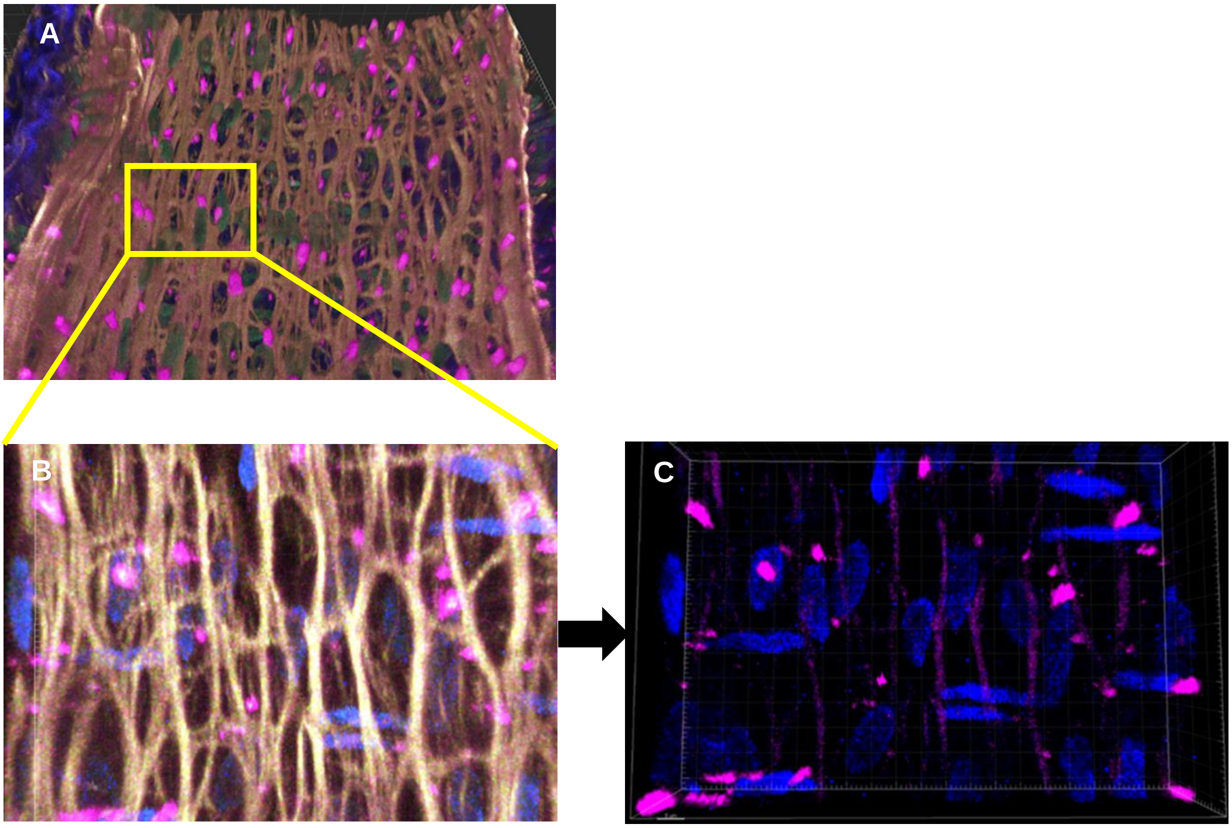


**D**

| | E.C. Nuclei | Globin Complexes | Complexes/Nuclei |
|---|---|---|---|
| ROI # 1 | 29 | 21 | 1.381 |
| ROI # 2 | 17 | 16 | 1.063 |
| ROI # 3 | 24 | 20 | 1.200 |

**Supplemental Figure 4 (A-D):** Representative schematic for calculating the ratio of hemoglobin-eNOS complexes to endothelial nuclei in an intact omental artery. **A)** A region of interest (ROI) is identified with in an image of an intact omental artery. **B)** The ROI is cropped to size, then **C)** the elastin autofluorescence is digitally subtracted to leave the autofluorescent complexes and DAPI-stained nuclei. The plane was then adjusted to exclude vascular smooth muscle nuclei (perpendicular alignment), and endothelial nuclei and complexes were counted in ImageJ. **D)** The total counts for three ROI from a single artery are then averaged to give a single ratio of complexes/nuclei.

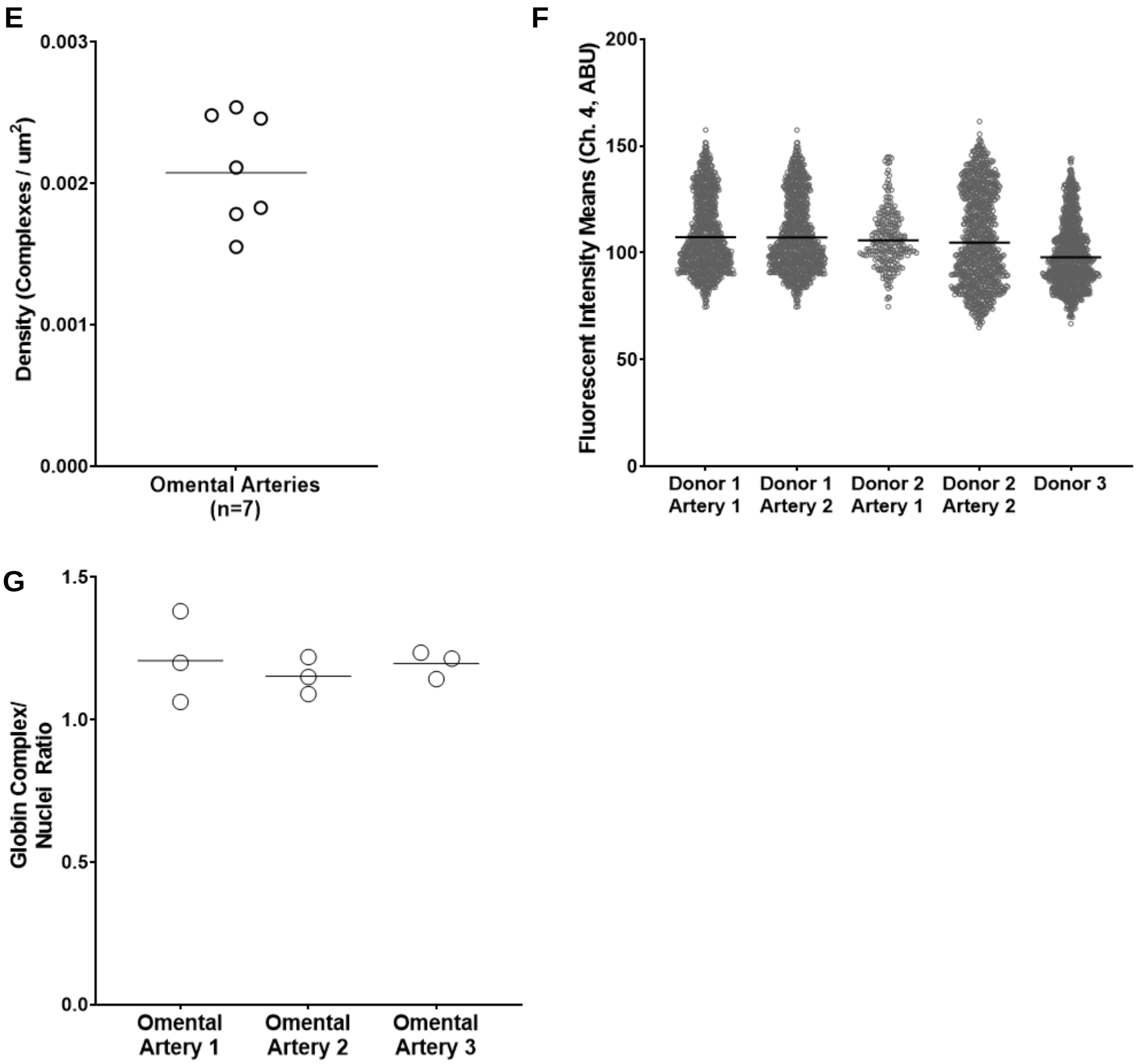


**Supplemental Figure 4 (E-G):** Spatial density, fluorescence intensity (Channel 4), and per nuclei ratio of hemoglobin-eNOS complexes in intact omental arteries. **E)**. To measure heterogeneity between arteries we calculated the density of punctates per analyzed surface area from seven donor arteries and found a consistent density of punctates. **F)** To measure heterogeneity between arteries from a single donor, and between donors, we analyzed the mean fluorescence intensity in channel 4 of hemoglobin-eNOS complexes from five total arteries (two from Donor 1, two from Donor 2, and one from Donor 3). Only the artery from Donor 3 was different and was slightly lower in mean channel 4 intensity. **G)** We calculated the ratio of globin complexes to nuclei as described in Supp. Fig. 5.

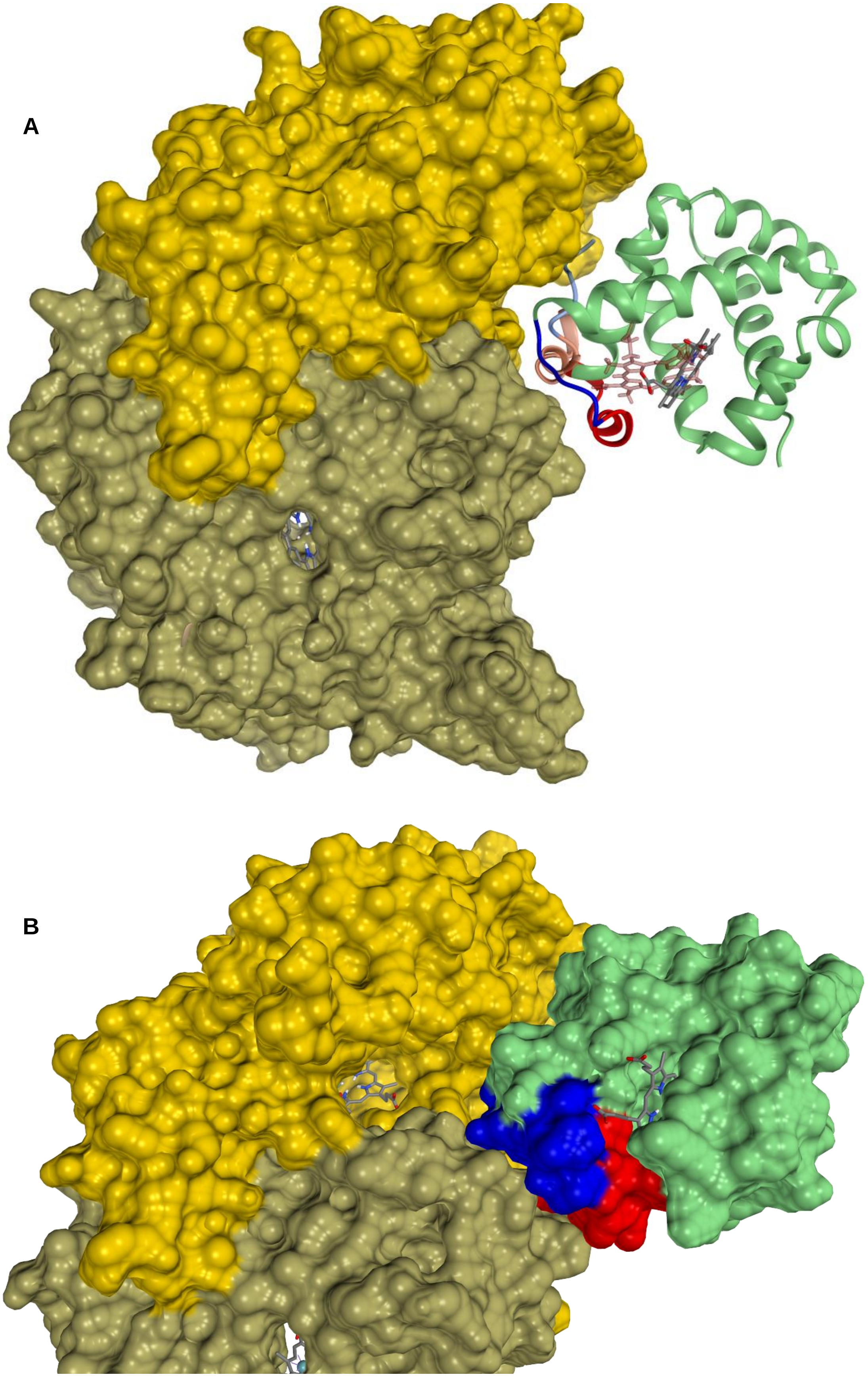

A
B

**Supplemental Figure 5: A)** Molecular modeling using UCSF Chimera based on computer docking of alpha globin (green ribbon on right of figure) with eNOS oxygenase domain homodimer (brown/gold indicating each chain). The alignment of alpha globin with eNOS was predicted using HADDOCK 2.4. The bright red ribbon denotes the peptide sequence of alpha globin that comprises the HbaX mimetic peptide, and the adjacent residues are shown in bright blue. For contrast, the predicted alignment of the HbaX peptide sequence with eNOS from Straub et al. 2014, originally published in ATVB, is shown in light red, with the adjacent residues shown in light blue. The new model shown here is shifted by several Ångstroms compared to the Straub et al. 2014 model, and allows for greater complimentary binding of full tetrameric hemoglobin with eNOS. **B)** Close-up view of the model showing alpha globin binding with eNOS; alpha globin has been shaded as a surface to show 3-D protein structure. The heme pocket of alpha globin and the heme pocket of eNOS are both visible, demonstrating their spatial proximity to each other to facilitate diffusion of NO from the eNOS reaction center to the alpha globin heme.

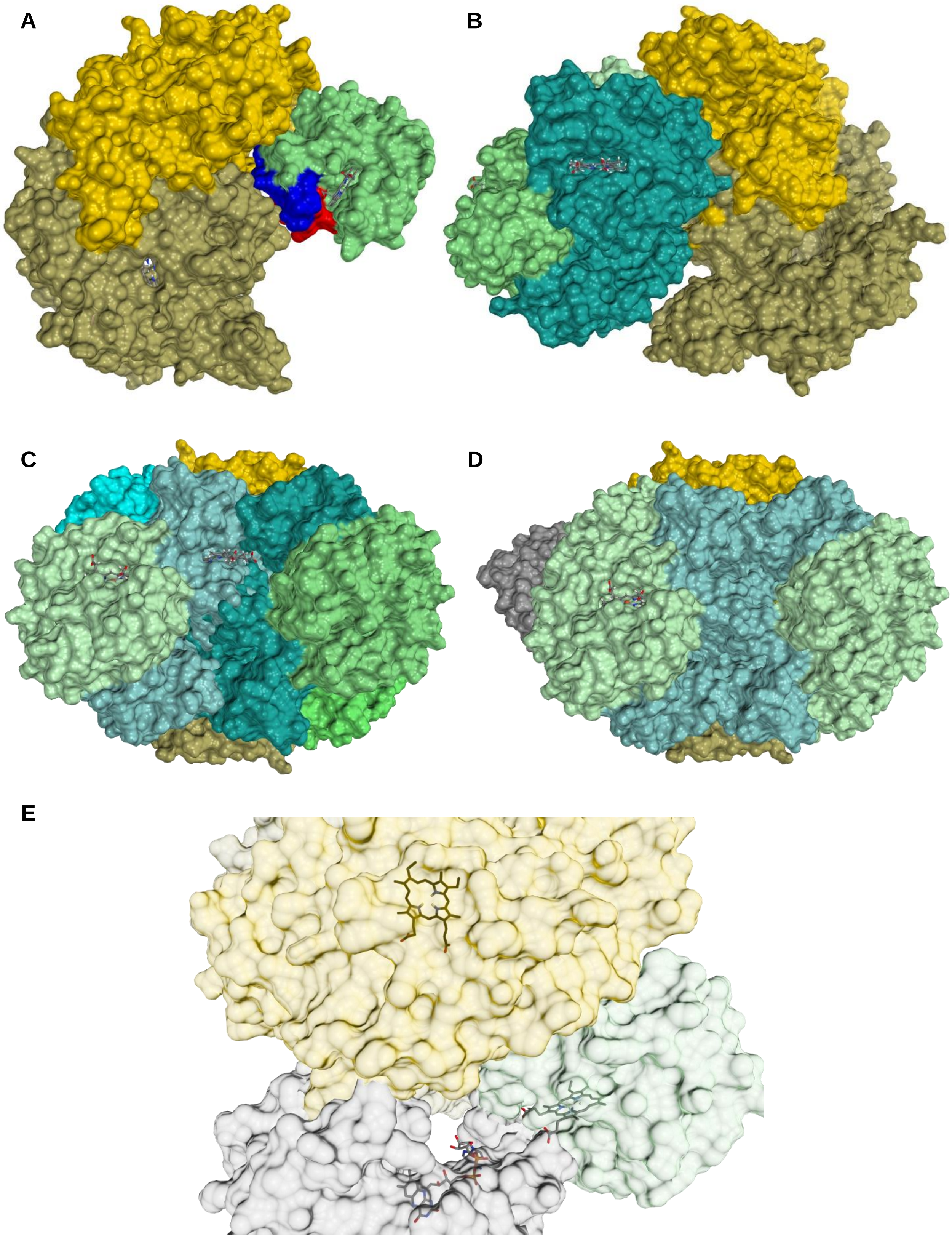
A
B
C
D
E

**Supplemental Figure 6:** Molecular modeling for alpha globin, tetrameric hemoglobin, Cyb5R3, and eNOS.(orange) aligned with eNOS (blue) created using UCSF Chimera, based on predictive docking models using HADDOCK 2.4. **A)** Predicted alignment for the alpha globin subunit of hemoglobin (green) with eNOS homodimer (gold/brown); the sequence of the HbaX mimetic peptide is shown in red with adjacent peptide residues in blue. **B)** Tetrameric hemoglobin aligned with eNOS- the original alpha globin monomer (light green, top of figure) remains in the same alignment as panel A; the second alpha subunit (green) and two beta globin units (teal) are shown in alignment with eNOS. Several peptide residues of beta globin are predicted to form stable bonds with residues of eNOS in this model, adding stability to the hemoglobin-eNOS complex. **C)** The model also accommodates the binding of a second hemoglobin with eNOS and the first hemoglobin tetramer- original hemoglobin is on left, with primary alpha subunit from panel A shown in bright blue, the second alpha globin in light green, and beta subunits in grey-blue. The second hemoglobin is on the right, with alpha subunits in brighter green and beta subunits in teal. **D)** Manual VR-enhanced docking of Cyb5R3 (grey) with alpha subunit of hemoglobin (light green) and eNOS (gold). **E)** VR-enhanced close-up of the predicted alignment between eNOS (gold), alpha globin (light green), and Cyb5R3 (grey) from the model in Panel D. The FAD pocket of Cyb5R3 is visible and is spatially very close with the heme pocket of alpha globin; the reduction of alpha globin by Cyb5R3 is an important mechanism in this model. Both Cyb5R3 and alpha globin bind with eNOS, with the eNOS heme pocket visible in close proximity as well.

**Supplemental Table 1:** Four parameter logistic regression best-fit values for artery pressure myography performed on pairs of omental arteries from 5 human donors.

| | **Top** | | **Bottom** | | **LogIC50** | | **HillSlope** | |
|---|---|---|---|---|---|---|---|---|
| **Mimetic Peptide (n = 5 human donors)** | **Mean** | **SEM** | **Mean** | **SEM** | **Mean** | **SEM** | **Mean** | **SEM** |
| **Phenylephrine (PE)** | 100.12 | 2.14 | 39.08 | 3.17 | -5.891 | 0.15 | -0.568 | 0.108 |
| **PE + Mimetic** | 99.93 | 0.83 | 64.59 | 1.55 | -5.575 | 0.12 | -0.549 | 0.076 |
| **PE + Mimetic + LNAME** | 100.58 | 1.19 | 41.89 | 2.02 | -5.768 | 0.10 | -0.497 | 0.054 |
| **Control Peptide (n = 5 human donors)** | **Mean** | **SEM** | **Mean** | **SEM** | **Mean** | **SEM** | **Mean** | **SEM** |
| **Phenylephrine (PE)** | 100.30 | 4.04 | 46.11 | 5.49 | -6.000 | 0.31 | -0.602 | 0.241 |
| **PE + Control** | 100.40 | 4.13 | 45.78 | 5.50 | -6.020 | 0.31 | -0.623 | 0.256 |
| **PE + Control + LNAME** | 100.40 | 3.36 | 45.40 | 4.55 | -6.021 | 0.25 | -0.582 | 0.187 |

**Supplemental Video 1 (see attached MP4 file):**

Image deconvolution and analysis was performed on the intact, perfused omental artery shown in Figure 5A using Imaris (Bitplane). This video shows the conversion of the raw image file into surface objects and then highlights anatomical features that can be viewed in the whole, intact omental artery.

1. (0:00- 0:08)- Raw image file of intact, perfused omental artery is converted into surface objects using Imaris (Bitplane). The artery and a small branch are visible. The outer collagen layer is rendered into a blue surface; the internal elastic lamina (IEL) is rendered into a yellow surface object; DAPI-labeled nuclei are rendered into light blue surface objects; hemoglobin-eNOS complexes are rendered into fuchsia surface objects. The view zooms in to a cropped subsection of the artery.
2. (0:08- 0:15)- The blue outer collagen surface is digitally removed, revealing the perpendicularly aligned vascular smooth muscle cell nuclei above the IEL.
3. (0:15- 0:20)- The IEL is subtracted digitally to show only the nuclei and the hemoglobin-eNOS complexes
4. (0:20- 0:40)- The IEL is added back, and the view is shifted to the axial plane. The top half of the artery is then digitally subtracted to enable a view of the artery from where the vessel lumen would be. The artery is then rotated back to be viewed from above, clearly showing endothelial nuclei in parallel with the direction of the IEL, and the hemoglobin-eNOS punctates in plane with the endothelial nuclei and the IEL.
5. (0:40- 0:45)- The IEL is made transparent, visualizing that the hemoglobin eNOS complexes are crossing and protruding through the IEL.
6. (0:46-0:58)- The IEL is returned to full color, and the artery rotated to be viewed from the side. The endothelial nuclei and smooth muscle nuclei are clearly visible, as are the hemoglobin eNOS complexes.
7. (0:58- 1:06)- The view is zoomed out and the collagen layer is added back, displaying the whole artery.
8. (1:06- 1:10)- The 3-D surfaces change back into the raw image file of the omental artery.

APPENDIX C- Multiphoton Microscopy Image Acquisition Settings

Data from: LAS X 3.5.6.21594

#### Dimensions

| Dimension | Logical Size | Physical Length | Start Position | End Position | Pixel Size / Voxel Size |
|---|---|---|---|---|---|
| X | 1024 | 387.5 µm | 0 µm | 387.5 µm | 0.379 µm |
| Y | 1024 | 387.5 µm | 0 µm | 387.5 µm | 0.379 µm |
| Z | 59 | 58.04 µm | 1,391.11 µm | 1,449.15 µm | 1.001 µm |

#### Channels

| LUT | Resolution | Min | Max | STED: DetectorMode / Huygens saturation factor / Wavelength |
|---|---|---|---|---|
| Blue | 8 | 0 | 255 | --- / --- / --- |
| Green | 8 | 0 | 255 | --- / --- / --- |
| Red | 8 | 0 | 255 | --- / --- / --- |
| Magenta | 8 | 0 | 255 | --- / --- / --- |

**Confocal Settings**

| Name | Value |
|---|---|
| Rotator Angle | -718.75 m° |
| Scan Mode | xyz |
| Scan Direction X | Bidirectional |
| Scan Speed | 600 Hz |
| Version Number | 15 |
| StagePosX | 7,323.59 µm |
| StagePosY | 41,665.99 µm |
| ZPosition | 1,449.15 µm |
| IsSuperZ | 0 |
| Magnification | 40 |
| ObjectiveName | HC PL IRAPO 40x/1.10 WATER |
| Immersion | WATER |
| Numerical Aperture | 1.1 |
| RefractionIndex | 1.33 |
| Zoom | 0.75 |
| Pinhole | 77.2 µm |
| PinholeAiry | 1 AU |
| EmissionWavelength for PinholeAiry Calculation | 580 nm |
| FrameAverage | 1 |
| LineAverage | 3 |
| FrameAccumulation | 1 |
| Line_Accumulation | 1 |
| IsUserSettingNameSet | 0 |
| IsRoiScanEnable | 0 |

### Filter Wheels / Other Motorized Devices

| Device Name | Filter Name/Position |
|---|---|
| RLD Blocking Slider | SP 800 |
| Merger Wheel | RSP 1005 |
| Galvo Slider | Galvo X Normal |
| Multi Function Port | SP 815 |
| Notch FW 2 | OPO SP 800 |
| Polarization FW | Empty |
| Galvo Resonant Pan | Galvo X Pan Center |
| Target Slider | Target Park |
| X2 Lens Changer | CS2 UV Optics 1 |
| VariableBeamExpander 1150nm | Factor: 1.03, Z Color Correction Offset: 0 |
| VariableBeamExpander 880nm | Factor: 1.03, Z Color Correction Offset: 0 |

### Lasers

| LaserName | OutputPower |
|---|---|
| **405 Diode** | **On** |
| **Argon** | **On, 19.8413 %** |
| **DPSS 561** | **On** |
| **HeNe 594** | **On** |
| **HeNe 633** | **On** |
| **Mai Tai DeepSee** | **On , Power: 2.2035 W** |
| **Insight** | **On , Power: 1.6000 W** |
| **Insight (Pump)** | **--** |

### Laser Lines

| Laser Line | Intensity |
|---|---|
| ( 405 nm) | Shutter: off, Intensity: 0.0000% |
| ( 458 nm) | Shutter: off, Intensity: 0.0000% |
| ( 476 nm) | Shutter: off, Intensity: 0.0000% |
| ( 488 nm) | Shutter: off, Intensity: 0.0000% |
| ( 496 nm) | Shutter: off, Intensity: 0.0000% |
| ( 514 nm) | Shutter: off, Intensity: 0.0000% |
| ( 561 nm) | Shutter: off, Intensity: 0.0000% |
| ( 594 nm) | Shutter: off, Intensity: 0.0000% |
| ( 633 nm) | Shutter: off, Intensity: 0.0000% |
| ( 880 nm) | Shutter: on, Intensity: 6.1281% |
| ( 1150 nm) | Shutter: on, Intensity: 7.3079% |

### Detectors

| Name | Channel | Type | Location | Active | Gain | Offset | Gate Start | Gate End | Gate Ref. Wavelength |
|---|---|---|---|---|---|---|---|---|---|
| PMT 1 | Channel 1 | PMT (401nm - 443nm) | Internal | Inactive | 0 | 0 | -- Time Gating not supported -- | | |
| HyD 2 | Channel 2 | HyD (485nm - 527nm) Standard mode | Internal | Inactive | 100 | -- | -- Time Gating not supported -- | | |
| PMT 3 | Channel 3 | PMT (569nm - 611nm) | Internal | Inactive | 0 | 0 | -- Time Gating not supported -- | | |
| HyD 4 | Channel 4 | HyD (653nm - 695nm) Standard mode | Internal | Inactive | 100 | -- | -- Time Gating not supported -- | | |
| HyD 5 | Channel 5 | HyD (737nm - 779nm) Standard mode | Internal | Inactive | 100 | -- | -- Time Gating not supported -- | | |
| PMT Trans | Transmission Channel | PMT | TLD | Inactive | 0 | 0 | -- Time Gating not supported -- | | |
| HyD-RLD1 | NDD1 | HyD (393nm - 472nm) Standard mode | RLD | Active | 170 | -0.01 | -- Time Gating not supported -- | | |
| HyD-RLD2 | NDD2 | HyD (496nm - 547nm) Standard mode | RLD | Active | 102.4 | -0.01 | -- Time Gating not supported -- | | |
| HyD-RLD3 | NDD3 | HyD (590nm - 621nm) Standard mode | RLD | Active | 100 | -0.01 | -- Time Gating not supported -- | | |
| HyD-RLD4 | NDD4 | HyD (658nm - 717nm) Standard mode | RLD | Active | 71 | -0.01 | -- Time Gating not supported -- | | |
